# Daily biofeedback to modulate heart rate oscillations affects structural volume in hippocampal subregions targeted by the locus coeruleus in older adults but not younger adults

**DOI:** 10.1101/2023.03.02.23286715

**Authors:** Hyun Joo Yoo, Kaoru Nashiro, Shubir Dutt, Jungwon Min, Christine Cho, Julian F. Thayer, Paul Lehrer, Catie Chang, Mara Mather

## Abstract

Using data from a clinical trial, we tested the hypothesis that daily sessions modulating heart rate oscillations affect older adults’ volume of a region-of-interest (ROI) comprised of adjacent hippocampal subregions with relatively strong locus coeruleus (LC) noradrenergic input. Younger and older adults were randomly assigned to one of two daily biofeedback practices for 5 weeks: 1) engage in slow-paced breathing to increase the amplitude of oscillations in heart rate at their breathing frequency (Osc+); 2) engage in self-selected strategies to decrease heart rate oscillations (Osc-). The interventions did not significantly affect younger adults’ hippocampal volume. Among older adults, the two conditions affected volume in the LC-targeted hippocampal ROI differentially as reflected in a significant condition x time-point interaction on ROI volume. These condition differences were driven by opposing changes in the two conditions (increased volume in Osc+ and decreased volume in Osc-) and were mediated by the degree of heart rate oscillation during training sessions.

## 1. Introduction

### 1.1. The locus coeruleus influences hippocampal function

The locus coeruleus (LC) is a small brainstem nucleus that provides most of the brain’s noradrenaline (Mather, 2020). In particular, the locus coeruleus (LC) densely innervates the hippocampus, especially the dentate gyrus, molecular layer, CA3, and CA4 subregions (Hagena et al., 2016; Hortnagl et al., 1991; Loy et al., 1980; Pickel et al., 1974). The LC modulates both long-term potentiation and long-term depression in the hippocampus (Hagena et al., 2016; Hansen, 2017; Nguyen and Gelinas, 2018), contributing to memory function. LC structure is associated with memory and cognition in aging (Dahl et al., 2019; Dutt et al., 2021; Wilson et al., 2013), especially with hippocampal-dependent episodic memory (Dahl et al., 2022).

Phasic rather than tonic modes of LC activity appear to benefit the hippocampus and memory function. Animal research indicates that LC neurons fire in two distinct modes. In one, on-going irregular but consistently high levels of LC neuron firing are associated with stress, higher distractibility, and poorer task performance, whereas in the other, LC neurons show only moderate tonic activity but fire in phasic bursts in response to salient stimuli and decisions (Aston-Jones et al., 1999; Vazey et al., 2018). Thus, during high tonic noradrenergic activity there is less available range for LC neurons to briefly increase their activity, limiting the ability to detect phasic noradrenergic responses (Aston-Jones and Cohen, 2005). When using optogenetic techniques to manipulate LC firing modes in rodents, phasic LC stimulation tends to have positive effects, enhancing memory and cognition, whereas tonic stimulation increases arousal and stress responses such as freezing (Ghosh et al., 2021; McCall et al., 2015; Takeuchi et al., 2016). Phasic LC activity contributes to hippocampal plasticity through activation of β-adrenergic receptors (Jurgens et al., 2005; O’Dell et al., 2010; Walling and Harley, 2004), which promote neurogenesis in the dentate gyrus (Bortolotto et al., 2019; Han et al., 2019; Jhaveri et al., 2010; Jhaveri et al., 2014). Experiments that inserted human pathological (hyperphosphorylated) tau into rat LC neurons indicate the tonic/phasic distinction is important in the context of tau pathology spreading from the LC to the medial temporal regions (Omoluabi et al., 2021). Daily phasic optogenetic stimulation of the LC in these rats prevented spatial and olfactory learning deficits whereas tonic stimulation of the LC did not (Omoluabi et al., 2021).

### 1.2. Breathing may lead the vagus nerve to modulate the LC in a phasic rather than tonic fashion

Existing research suggests that vagus nerve activity promotes phasic LC activity. Afferent vagus nerve fibers project to the nucleus tractus solitarius (NTS), which in turn projects to the LC (Ruffoli et al., 2011). In rodents, stimulating the vagus nerve drives rapid, phasic neural activity in the LC (Hulsey et al., 2017). Importantly, the predominant physiological drivers of vagus nerve traffic should stimulate intermittent rather than steady LC activity. For instance, the vagus nerve participates in a negative-feedback neural loop that regulates blood pressure (Kaufmann et al., 2020). Baroreceptors are mechanical receptors that sense stretch in blood vessels (mainly the carotid arteries and aorta) and send signals via the vagus nerve (Stocker et al., 2019). These signals are relayed from the NTS to the dorsal motor nucleus that triggers efferent signals to the heart to slow down, to reduce blood pressure (Ruffoli et al., 2011).

Because these signals take a few seconds to complete the feedback loop, the feedback is always a bit out of date, leading blood pressure to continue to increase while ‘decrease’ signals are being sent (overshooting the target zone), and then, once reversed, to decrease even after ‘decrease’ signals are no longer being sent (deBoer et al., 1987; Vaschillo et al., 2006). The approximately 5-s delay leads to a ∼10-s period of increasing and then decreasing heart rate, or a spectral frequency peak in heart rate at around 0.1 Hz (Vaschillo et al., 2006). Thus, baroreceptors provide sensory signals to the vagus nerve in a regular periodic rhythm, which in turn transmits signals to the LC via the NTS (Ruffoli et al., 2011).

Breathing also is associated with phasic vagus nerve signals (Dergacheva et al., 2010). Recording of single-neuron activity within rats’ vagus nerve revealed burst activity corresponding with breathing frequency (Jiman et al., 2020), which may reflect signals from stretch receptors in the lungs (Schelegle and Green, 2001) or other signals from the body that differ in intensity during inhalation vs. exhalation, such as input from baroreceptors or chemoreceptors (Chapleau and Sabharwal, 2011). These breathing-related variations in body signals should in turn modulate vagus-nerve-to-LC signaling. Furthermore, vagus nerve signals are transmitted to the LC more effectively when exhaling than inhaling (Sclocco et al., 2019), potentially due to inhibition of the nucleus tractus solitarius when inhaling (Miyazaki et al., 1999). Consistent with the notion that breathing biases LC activity against a steady tonic pattern, functional magnetic resonance imaging suggests that, during rest, locus coeruleus activity fluctuates with breathing (Melnychuk et al., 2018), and in an in-vitro brainstem-spinal cord preparation of the neonatal rat, most locus coeruleus neurons’ activity was modulated at the respiratory frequency (Oyamada et al., 1998).

### 1.3. Effects of vagus nerve activity on hippocampal-dependent memory processes

Via its modulation of the LC-noradrenergic system, the vagus nerve can influence hippocampal function (Raedt et al., 2011; Roosevelt et al., 2006). Studies in rats reveal that pulses of vagus nerve stimulation affects both hippocampal plasticity and memory performance. For example, a 3-hr session of intermittent pulses of vagus nerve stimulation increased markers of neurogenesis (BrdU^+^ cells and doublecortin expression) in the dentate gyrus (Biggio et al., 2009), phasic (30 s on, 5 min off) pulsed vagus nerve stimulation at 0.75 mA promoted granule cell proliferation in the hippocampus (Revesz et al., 2008), one 30-s set of pulses of vagus nerve stimulation after an inhibitory-avoidance task improved avoidance memory a day later (Clark et al., 1998) and 30 min of intermittent pulses of vagus nerve stimulation during object familiarization improved next-day novel object preference (Sanders et al., 2019). Rats receiving a brain injury showed better Morris water maze performance two weeks later if they were receiving 30 min/day of vagus nerve stimulation (30.0-sec trains of 0.5 mA, 20.0 Hz, biphasic pulses) than if they received no vagus nerve stimulation (Smith et al., 2005). In another study with rats, ablating sensory afferent vagus nerve transmission impaired hippocampal-dependent memory and decreased markers of neurogenesis (Suarez et al., 2018), revealing that vagus afferent signaling affects the hippocampus not just during artificial vagus nerve stimulation, but also at endogenous levels of vagus nerve activity.

### 1.4. Hypothesized benefit of resonance frequency breathing for the aging hippocampus

As outlined in Section 1.2, both the baroreflex and breathing produce oscillations in vagus nerve inputs to the LC. Usually people breathe at a faster frequency (around 0.25 Hz or 4-s/breath) than the ∼0.1 Hz (10-s) baroreflex oscillatory influence. However, if people slow their breathing to a ∼10-s rhythm (5-s inhale, 5-s exhale), the two signals can influence heart rate at the same frequency, creating resonance (Lehrer and Gevirtz, 2014; Vaschillo et al., 2006). Furthermore, research comparing pharmacological sympathetic vs. parasympathetic blockade indicates that increases in spectral power at the breathing frequency during slow paced breathing are almost entirely vagally mediated (Kromenacker et al., 2018). Thus, via its stimulation of these vagus-LC pathways, slow breathing should induce bursts of noradrenergic release throughout the extended LC efferent network, especially in those subregions of the hippocampus that are densely innervated by the LC. At the same time, given the inhibitory synapses also present in the vagus-LC pathways (Fornai et al., 2011), resonance frequency breathing may lead inhibitory signals to alternate with the excitatory signals. This strong oscillating vagus nerve signal could help regulate the aging LC, promoting a beneficial phasic burst activity pattern (Omoluabi et al., 2021), in contrast with the high tonic levels of noradrenergic activity that increasingly appear to be the default as people age (see Mather, 2021 and next section).

### 1.5. Aging is associated with tonic noradrenergic hyperactivity

In humans, cerebrospinal noradrenaline and its metabolites increase with age and Alzheimer’s disease (for reviews see Mather, 2021; Weinshenker, 2018). Salivary alpha amylase provides a non-invasive proxy measure of central norepinephrine activity (Warren et al., 2017) and salivary alpha amylase levels also are greater in older than younger (Almela et al., 2011; Birditt et al., 2018; Nater et al., 2013; Strahler et al., 2010). Compared with younger mice, older mice show up-regulated proteins related to synaptic activity and other aspects of metabolic activity in the LC (Evans et al., 2021). Thus, evidence suggests hyperactive noradrenergic activity in the brain in aging.

EEG and pupil measures suggest that older adults have lower phasic LC responses than younger adults. Older adults and Alzheimer’s disease patients show reduced amplitude and increased latency of P3 responses to oddball stimuli (e.g., Polich and Corey-Bloom, 2005; van Dinteren et al., 2014; Walhovd and Fjell, 2001). Because phasic LC activity facilitates the P3 event-related potential (Nieuwenhuis et al., 2005; Vazey et al., 2018), older adults’ impaired P3 could reflect lower phasic LC activity, although it could alternatively reflect other factors such as age-related changes in cortical regions associated with the P3 (Mendes et al., 2022). In addition, a latent measure of noradrenergic responsiveness that included pupil dilation and EEG measures was lower in older than younger adults (Dahl et al., 2020).

### 1.6. Overview of current study

Based on the findings reviewed above, we hypothesized that heart rate oscillation biofeedback involving slow-paced breathing could affect older adults’ hippocampal volume, especially in hippocampal subregions regions with strong noradrenergic innervation. We used data from a recently completed clinical trial of heart rate oscillation biofeedback for this purpose (ClinicalTrials.gov NCT03458910; Heart Rate Variability and Emotion Regulation or “HRV-ER”). Data associated with this clinical trial are publicly available and we have provided a corresponding data description paper (Yoo et al., 2022b). The pre-registered outcome measures in this trial focused on changes in emotion-related brain networks (see Nashiro et al., 2023). Based on prior correlations between individual differences in left orbitofrontal cortex structure and HRV (Koenig et al., 2021; Yoo et al., 2018), we also investigated the intervention effects on structural volume in left orbitofrontal cortex regions (Yoo et al., 2022a). Hippocampal structural change is an exploratory endpoint of interest given the hypothesized effects of the intervention on the LC outlined above. In this trial, 106 younger and 56 older participants completed five weeks of daily heart rate oscillation biofeedback. The two intervention conditions both included twice-daily practice sessions that were 10-20 min long involving a continuous stream of biofeedback about their current heart rate with a rolling window of about three minutes of the heart rate history shown. However, participants in the two conditions had opposite objectives. In the Increase-Oscillations (Osc+) condition, participants were instructed to increase the amplitude of breathing-induced heart rate oscillations by breathing slowly in rhythm with a visual pacer. In contrast, in the Decrease-Oscillations (Osc-) condition, participants were instructed to try to keep their heart rate steady using self-selected strategies, such as imagining the ocean, listening to nature sounds, or listening to instrumental music. We conducted magnetic resonance imaging (MRI) structural scans of participants’ brains both before and after the intervention.

To form our hippocampal region-of-interest, we used Freesurfer to segment subregions of the hippocampus. We focused on subregions within the hippocampal body as there is more agreement on standards for segmenting the body than the head or tail (Bender et al., 2018; Olsen et al., 2019). Within the hippocampal body, we aggregated across right and left subregions for which there is evidence that they are densely or moderately innervated by the LC, namely the molecular layer, CA3, CA4 and granule cell layer of dentate gyrus (Hagena et al., 2016; Hortnagl et al., 1991; Loy et al., 1980; Pickel et al., 1974).

As our intervention could affect cerebrovascular dynamics, which in turn could potentially affect brain volume, we also examined whether there were changes in cerebral blood flow (CBF) within the hippocampal region of interest, using pulsed continuous arterial spin labeling (pCASL; Chen et al., 2011).

## 2. Materials and methods

### 2.1. Participants

We recruited 121 younger participants (aged 18-35 years) and 72 older participants (aged 55-80 years) via the USC Healthy Minds community subject pool, a USC online bulletin board, Facebook and flyers. Prospective participants were screened for medical, neurological, or psychiatric illnesses. We excluded people who had a disorder that would impede performing the heart rate oscillation biofeedback procedures (e.g., coronary artery disease, angina, cardiac pacemaker), who were currently practicing relaxation, biofeedback or breathing techniques, or were on any psychoactive drugs other than antidepressants or anti-anxiety medications. We included people who were taking antidepressant or anti-anxiety medication and/or attending psychotherapy only if the treatment had been ongoing and unchanged for at least three months and no changes were anticipated (see Supplementary Table S1 for categories of medications participants were taking). For older adults, we also excluded people who scored lower than 16 on the TELE (Gatz et al., 1995) for possible dementia. Gender, education, age and race were similar in the two conditions.

Participants provided written informed consent approved by the University of Southern California (USC) Institutional Review Board. Participants were recruited in waves of approximately 20 participants from the same age group, each of whom was assigned to small groups of 3-6 people, with each group meeting at the same time and day each week. After recruitment and scheduling of each wave of groups were complete, we assigned each group to one of two conditions involving daily biofeedback that aimed to increase heart rate oscillations (Osc+ condition) or to not increase heart rate oscillations despite a relaxed state (Osc-condition). We used randomization across small groups to maintain balanced numbers of each condition. We did this by determining how many groups were assigned to a condition; for example, if 2 out of 5 groups in a previous wave were assigned the Osc+ condition, then 3 out of 5 groups were assigned that condition in the next wave. Then we randomly allocated those conditions to groups. Younger and older adults were run in separate waves of groups, thus participants were assigned to groups with other participants from the same age group. One research staff member who was blinded to participants and small group assignment generated the random numbers and assigned the conditions to each small group. The study utilized a single-blinded design; the consent document did not mention that there were two conditions. Among enrolled participants, 12 participants dropped out before the beginning of the intervention, leaving 117 younger adults (N_Osc+_ = 60, N_Osc-_ = 57) and 64 older adults (N_Osc+_ = 33, N_Osc-_ = 31) who remained at the beginning of the intervention at week 2 (see Supplementary Figure S1 for flow diagram). During the intervention, 19 participants (N_Osc+_ = 9, N_Osc-_ =10) dropped out. 106 younger adults (N_Osc+_ = 56, N_Osc-_ = 50) and 56 older adults (N_Osc+_ = 28, N_Osc-_ = 28) completed all 7 weeks of study. For each analysis using each measure, we used the maximal number of participants available after excluding outliers (see Supplementary Figure S1 for flow diagram).

### 2.2. Procedure

#### 2.2.1. Overview of 7-week protocol schedule

The study protocol involved seven weekly lab visits and five weeks of home biofeedback training (see Supplementary Figure S2 for flow diagram). The first lab visit involved the non-MRI baseline measurements. The second lab visit involved the baseline MRI session, followed by the first biofeedback training session. Each lab biofeedback training session started with a 5-min baseline rest period and individual calibration to find the best training condition. During the second lab visit participants were provided with 11.6-inch laptops with the biofeedback software and ear sensor units for lab sessions and training at home. The final (7th) lab visit repeated the baseline MRI session scans in the same order (for more details see Supplementary Materials or our data description paper for the clinical trial; Yoo et al., 2022b).

#### 2.2.2. Biofeedback Training

During all practice sessions, participants wore an ear sensor to measure their pulse. In the Osc+ condition, participants viewed real-time heart rate biofeedback while breathing in through the nose and out through the mouth in synchrony with the emWave pacer. The emWave software (HeartMath®Institute, 2020) provided a summary ‘coherence’ score (see Supplementary Materials for more details). During calibration, the resonance frequency was determined individually. Participants in Osc+ condition were then instructed to train at home with the pacer set to their identified resonance frequency and to try to maximize their coherence scores (see Fig. 1A, C and Figure S3A).

**Fig. 1.**
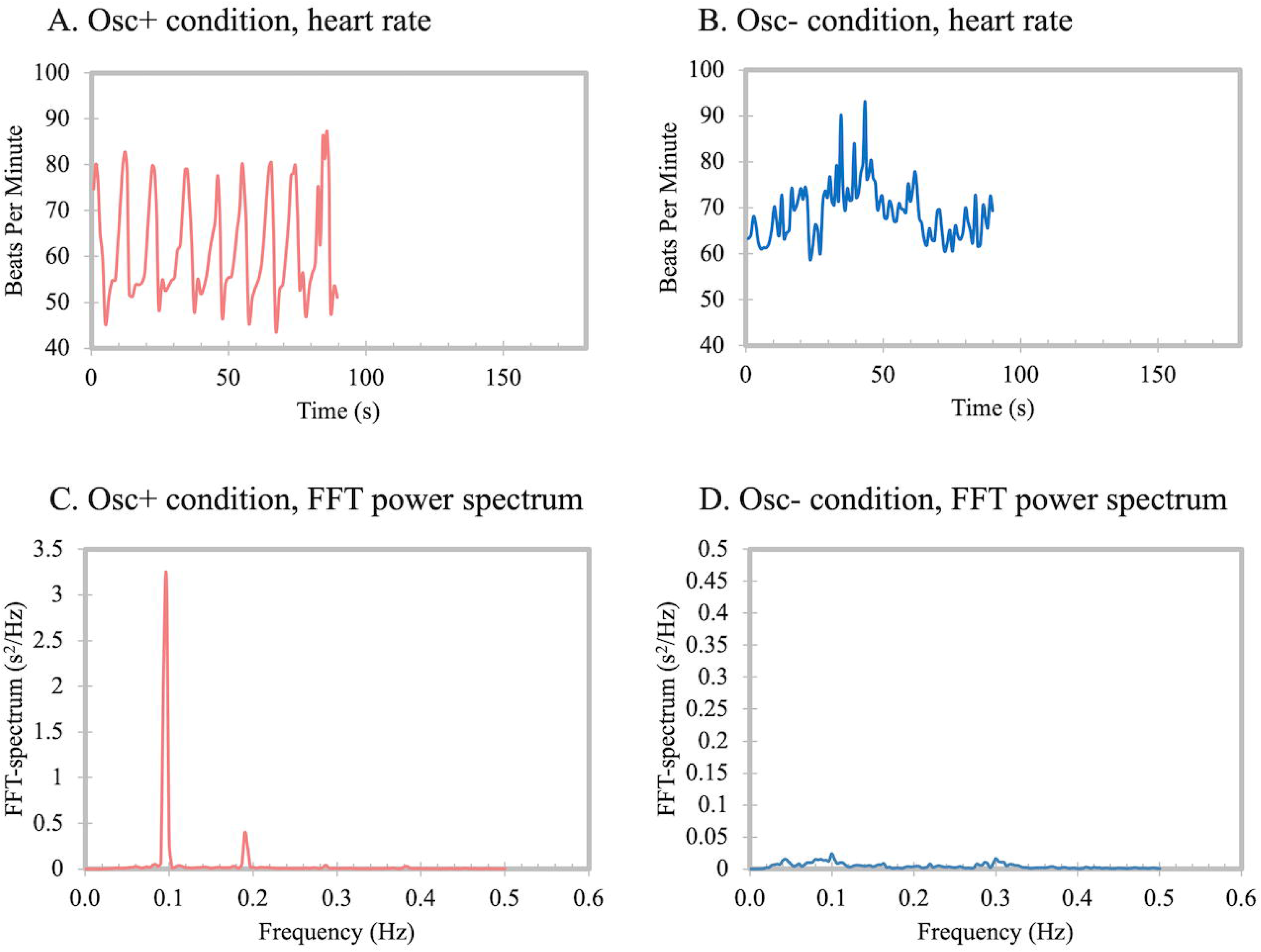
Examples of biofeedback display for Osc+ condition (A) and Osc-condition (B) and examples of power spectrum using fast Fourier transform (FFT) method for Osc+ condition (C) and Osc-condition (D). Note that in order to allow Osc+ power spectrum to be visible, the Y axis scales differ in panels C and D. In the Increase-Oscillations (Osc+) condition, participants were instructed to maximize heart rate oscillatory activity by breathing slowly in rhythm with a visual pacer. In the Decrease-Oscillations (Osc-) condition, participants were instructed to try to keep their heart rate steady using self-selected strategies.

In the Osc-condition, the same biofeedback ear sensor was used. However, custom software displayed a different set of feedback to the Osc-participants. During each Osc-training session, a ‘calmness’ score provided feedback to the participants instead of the coherence score (see Supplementary Materials for more details). Participants in Osc-condition were then instructed to train at home with their most effective strategy and to try to lower heart rate and heart rate oscillations (see Fig. 1B, D and Figure S3B).

Upon completing the study, participants were paid for their participation and received bonus payments based on their individual and group performances. As outlined in Nashiro et al., (2023), participants were assigned personalized targets (their top 10 biofeedback coherence or calmness scores earned from the previous week’s training sessions averaged, plus 0.3) each week. Individual bonus amounts were calculated for each week based on how many training sessions they exceeded their assigned target score and a group bonus was added for each week when members of a participant’s group completed a minimum of 80% of their training minutes (incentives for training were the same across conditions; see details in the Supplementary Methods under “Rewards for Performance” and Supplementary Table S2).

### 2.3. MRI scan parameters

The scans were conducted with a 3T Siemens MAGNETOM Trio scanner with a 32-channel head array coil at the USC Dana and David Dornsife Neuroimaging Center. T1-weighted 3D structural MRI brain scans were acquired pre- and post-intervention using a magnetization prepared rapid acquisition gradient echo (MPRAGE) sequence with the parameters of TR = 2300 ms, TE = 2.26 ms, slice thickness = 1.0 mm, flip angle = 9°, field of view = 256 mm, and voxel size = 1.0 x 1.0 x 1.0 mm, with 175 volumes collected (4:44 min). Prior to the T1-weighted structural scans, pseudo-continuous arterial spin labeling (pCASL) scans were acquired with TR = 3880, TE = 36.48, slice thickness = 3.0 mm, flip angle = 120°, field of view = 240 mm and voxel size = 2.5 × 2.5 × 3.0 mm, with 12 volumes collected (3:14 min; 1^st^ volume was an M0 image, 2^nd^ volume was a dummy image, and the remaining 10 volumes were 5 tag-control pairs) during resting-state. This ASL approach provides high precision and signal-to-noise properties and has better test-retest reliability than pulsed or continuous ASL techniques (Chen et al., 2011).

### 2.4. Analyses

#### 2.4.1. T1-Weighted neuroimaging processing

Among the 106 younger adults and 56 older adults who completed all 7 weeks of the study, 100 younger adults and 51 older adults finished both pre- and post-intervention MRI sessions. Each participant’s T1 structural image was preprocessed using Freesurfer image analysis suite version 6.0 (http://surfer.nmr.mgh.harvard.edu/). Cortical reconstruction and volumetric segmentation were performed. This method uses both intensity and continuity information from the entire three-dimensional MR volume in segmentation and deformation procedures to produce representations of cortical thickness, calculated as the closest distance from the gray/white boundary to the gray/CSF boundary at each vertex on the tessellated surface (Fischl and Dale, 2000). The technical details of these procedures are described in prior publications (Fischl et al., 2002; Fischl et al., 2004a; Fischl et al., 2004b).

After basic processing, we used the Freesurfer 6.0 image analysis suite longitudinal stream to automatically extract volume estimates (Reuter et al., 2012). This longitudinal stream within FreeSurfer creates an unbiased subject-specific template created by co-registering scans from the pre- and post-intervention time-points using a robust and inverse consistent algorithm (Reuter et al., 2010; Reuter et al., 2012), and uses these subject-specific templates for pre-processing the individual scans (e.g., during skull stripping and atlas registration). This approach improves reliability and statistical power while avoiding processing bias favoring the baseline scans that may affect groups unequally (Reuter et al., 2012).

#### 2.4.2. Automatic segmentation of hippocampal subfields

We employed FreeSurfer v6.0’s segmentation tool of hippocampal subfields and nuclei of the amygdala for longitudinal segmentation (https://surfer.nmr.mgh.harvard.edu/fswiki/HippocampalSubfieldsAndNucleiOfAmygdala). The algorithm for segmentation of individual subregions uses Bayesian inference based on observed image intensities and a probabilistic atlas built from a library of in vivo manual segmentations and ultra-high resolution (∼0.1 mm isotropic) ex vivo labeled MRI data (Iglesias et al., 2015; Iglesias et al., 2016)). The hippocampus and amygdala were jointly segmented to avoid overlap or gaps between structures. The longitudinal pipeline uses a binary mask of the hippocampus extracted from the automated segmentation of each subject’s base template segmented 19 hippocampal subfields.

For quality control, we used automated measures computed by FreeSurfer of the contrast-to-noise ratio (the difference in signal intensity between regions of different tissue types and noise signal) and the Euler number (a metric of cortical surface reconstruction) to identify poor quality structural scans (Chalavi et al., 2012; Rosen et al., 2018). We excluded outliers (N=4 for younger adults and N=2 for older adults) who on a box-and-whisker plot were above Q3 + 3 * the interquartile range on either of these metrics on either pre-intervention or post-intervention scans. One older adult was excluded due to the failure to segment the nuclei of the amygdala and the hippocampal subfields in the right hemisphere. We followed the quality control procedure guidelines for the FreeSurfer-based segmentation of the hippocampal subregions (Samann et al., 2022). First, we checked for outliers (+/− 2 SDs, representing roughly 5% of cases in the distribution of our dataset) for each subfield volume, and also for total brain volume, total GM volume, ICV, and GM/ICV ratio. Second, the deviation pattern of the rank order of subfield volumes was calculated within a subject based on the rank order from several large datasets. In addition, the histograms of each measure were inspected. As a final step, we visually inspected all datasets in the HTML files with a standard browser, with particular attention to the flagged datasets from the previous steps. We followed the guideline for visual QC; 1) Is binary hippocampal mask visible? 2) Is hippocampal fissure positioned within hippocampal mask? 3) Are larger portions of the hippocampus cut off? 4) Are there any subfield-related peculiarities? There was no additional exclusion after this QC pipeline. No manual edits were carried out to avoid the introduction of external bias. The final hippocampal volume dataset had an N of 96 for younger adults (N_Osc+_ = 49; N_Osc-_ = 47) and an N of 48 for older adults (N_Osc+_ = 23; N_Osc-_ = 25).

The ROI included eight subregions densely innervated by the LC; left and right molecular layer, CA3, CA4 and granule cell layer of dentate gyrus. Volumes were extracted for each of the eight subregions and summed as ROI volume. In addition, the volume of the whole hippocampus was also analyzed for comparison.

#### 2.4.3. Heart rate oscillations during seated rest

Pulse signal was acquired using the emWave ear sensor. The emWave ear sensor has been evaluated as a reliable and low-cost wearable device (Lo et al., 2017). To measure and compare heart rate and HRV consistently, we used participants’ home training device during rest at the lab as well as during home training. To assess whether the interventions affected heart rate oscillatory activity during rest, we used Kubios HRV Premium 3.1 (Tarvainen et al., 2014) to compute heart rate and the main HRV indices (heart rate, SDNN, RMSSD, LF power and HF power, total power using autoregressive method during the baseline rest sessions (5-min sessions before lab training sessions) in the lab in weeks 2 vs. 7.

In younger adults, we excluded outliers (N=5) who on a box-and-whisker plot were above Q3 + 3 * the interquartile range on total heart rate spectral power for pre-intervention rest (N=3), post-intervention rest (N=1), or average training (N=1), leaving an N of 97 for younger adults (N Osc+ = 52; N Osc-= 45) and in older adults, we excluded outliers (N=3) who on a box-and-whisker plot were above Q3 + 3 * the interquartile range on total power for pre-intervention rest (N=1), or post-intervention rest (N=2), leaving an N of 53 for older adults (N Osc+ = 25; N Osc-= 28). The common dataset from pre/post-intervention heart rate, HRV and MRI data had an N of 88 for younger adults (N_Osc+_ = 45; N_Osc-_ = 43) and an N of 46 for older adults (N_Osc+_ = 21; N_Osc-_ = 25).

Before conducting statistical analyses, the Shapiro–Wilk test was run for heart rate and HRV metrics to ensure normal distribution. All these metrics except mean HR were not normally distributed (p_<_0.05). To correct for this, SDNN, RMSSD, HF power, LF power, and total power were transformed using the natural log function.

#### 2.4.4. Heart rate oscillations during training and its relationship with post-intervention hippocampal ROI volume

To assess the impact of Osc+ versus Osc-biofeedback during training sessions, we used Kubios HRV Premium 3.1 (Tarvainen et al., 2014) to compute autoregressive spectral power for each training session. We analyzed data from the same set of participants as for resting HRV (see previous section), 88 younger adults (M = 56.84, SE = 2.2 sessions) and 46 older adults (M = 78.52, SE = 2.5 sessions). We extracted the summed power within the .063∼.125 Hz range for each participant to obtain a measure of heart rate oscillatory activity during biofeedback. We used the frequency range corresponding with 8-16s as it encompassed the resonance frequency paces used by Osc+ participants for their breathing and thereby allowed us to compare differences in heart rate oscillatory activity in this range influenced by slow breathing (between 8 and 15.87 s) and the baroreflex (Kuusela et al., 2003) across the two conditions. Before conducting statistical analyses, we log transformed the power values. We conducted partial correlations of these estimates of resonance frequency power during training with the post-intervention hippocampal ROI, controlling for the pre-intervention ROI. This partial correlation approach of controlling for the baseline ROI value is akin to using a pre-intervention - post-intervention difference score of the hippocampal ROI while first equating participants’ baseline scores (Burt and Obradović, 2013). Thus, it reflects the relationship between change in the hippocampal ROI volume and heart rate oscillations during training while controlling for baseline differences in hippocampal ROI volume.

To examine whether the relationships between the training condition and the ROI volume changes were mediated by the resonance frequency power within the .063∼.125 Hz range during training, we conducted a mediation analysis using the PROCESS macro version 4.2 (Hayes, 2017). In the model, we included the condition as an independent variable, resonance frequency power during training as a mediator, and the ROI volume at post-intervention as a dependent variable. We included the pre-intervention ROI volume as a control variable, such that the model addressed the changes in ROI volume. The unstandardized regression coefficient (c) reflects the total effect. Coefficient c′ reflects the direct effect of the independent variable on the dependent variable absent the mediator. Coefficients a and b reflect the relationships between the mediator and the independent variable and the dependent variable, respectively. The product of coefficients (a × b) indicates how much the relationship between the independent variable and dependent variable is mediated by the mediator (i.e., the indirect effect).

#### 2.4.5. Arterial Spin Labeling

Data were preprocessed using the Arterial Spin Labeling Perfusion MRI Signal Processing Toolbox (ASLtbx; Wang et al., 2008). M0 calibration image and 10 tag-control pairs were motion corrected, co-registered to individual participants’ T1-weighted structural images, smoothed with a 6 mm full width at half maximum Gaussian kernel, and normalized to MNI template space. Preprocessing resulted in a time-series of 5 perfusion images representing the tag-control pairs, which were averaged to create a single mean whole brain perfusion image. We included a study-specific gray matter mask comprised of averaged gray matter segmentations across participants’ T1-weighted structural scans in all voxel-wise analyses to restrict analyses to gray matter CBF, as ASL has lower power to detect white matter than gray matter perfusion signal (van Osch et al., 2009). Images were thresholded at 0 ml/100g/min to remove background voxels and voxels with negative values (Bangen et al., 2014; Brown et al., 2003).

To apply ROI masks on CBF images, FreeSurfer’s labeled hippocampus segmentations were converted back to the native MR image space using the procedure provided by FreeSurfer (mri_label2vol), and linearly transformed into MNI152 2 mm standard space. This process was carried out for the left and the right hippocampus separately. Averaged CBF values from whole brain gray matter regions and from hippocampal ROI were calculated.

Among 144 participants with hippocampal volume data, 84 younger adults (N_Osc+_ = 42; N_Osc-_ = 42) and 47 older (N_Osc+_ = 22; N_Osc-_ = 25) completed pCASL data at pre- and post-intervention sessions. 26 younger adults (N_Osc+_ = 14; N_Osc-_ = 12) and 18 older adults (N_Osc+_ = 8; N_Osc-_ = 10) were excluded due to errors in preprocessing or excessive motion, resulting in a total of 58 younger adults (N_Osc+_ = 28; N_Osc-_ = 30) and 29 older adults (N_Osc+_ = 14; N_Osc-_ = 15) in subsequent pCASL analyses involving pre- and post-intervention scans.

## 3. Results

### 3.1. Sample characteristics

Details about the demographic characteristics of the participants can be found in Table 1. Participants’ daily medication reports are shown in Supplementary Table S1.

**Table 1.**
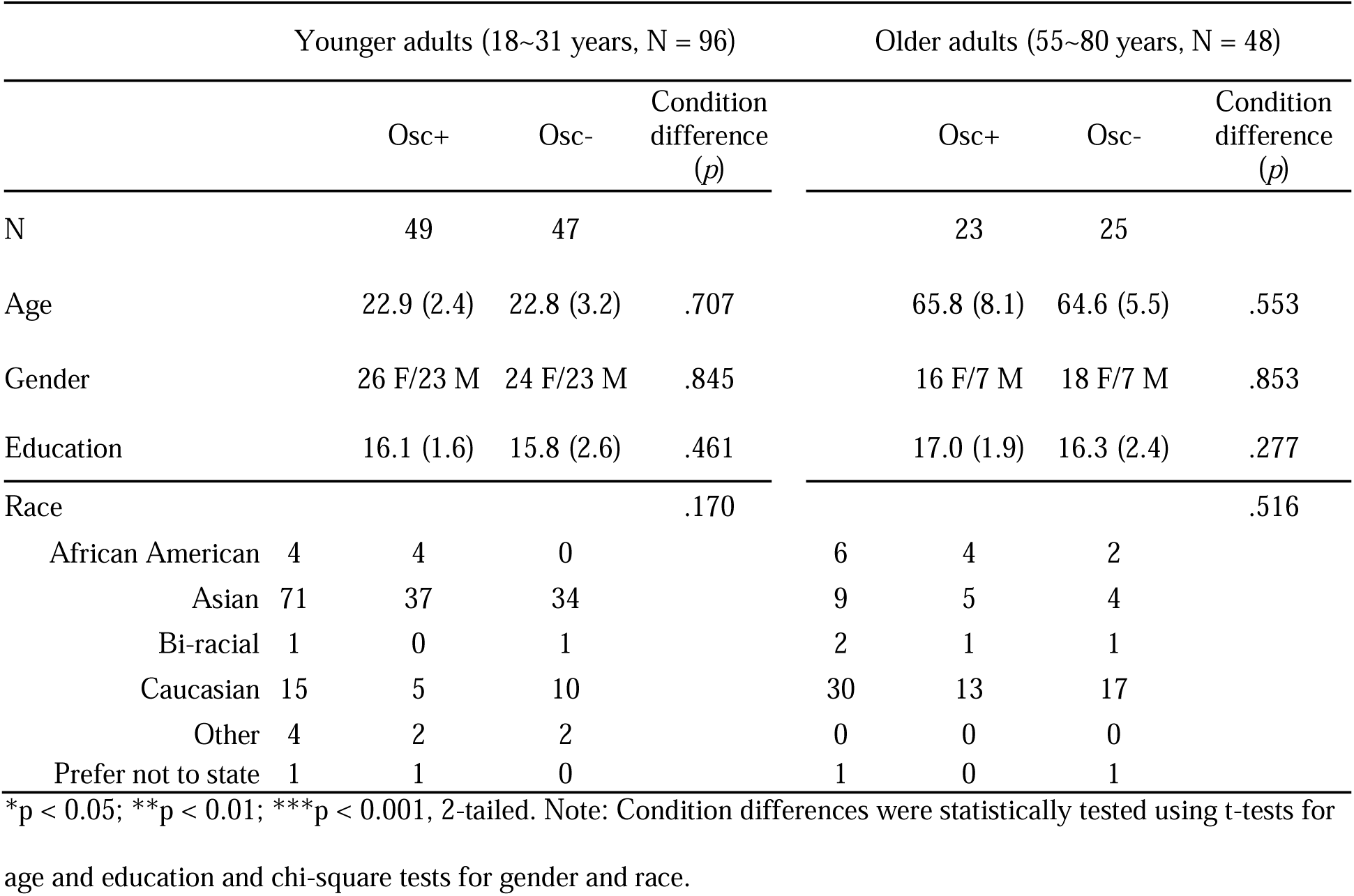
Basic participant characteristics across age groups and conditions (N=144)

### 3.2. Volume in the LC-innervated-regions hippocampal ROI

The hippocampal subfields and nuclei of the amygdala segmentation algorithm of the FreeSurfer v6.0 was applied to T1 weighted images according to the longitudinal pipeline (Iglesias et al., 2016). In Fig. 2, axial, coronal, and sagittal views of hippocampal subfields segmentation from an older adult participant enrolled in the study are shown. Table S3 in the supplementary information summarizes the list of hippocampal subfield segmentation and the test–retest reliabilities of the subfield volume estimation.

**Fig. 2.**
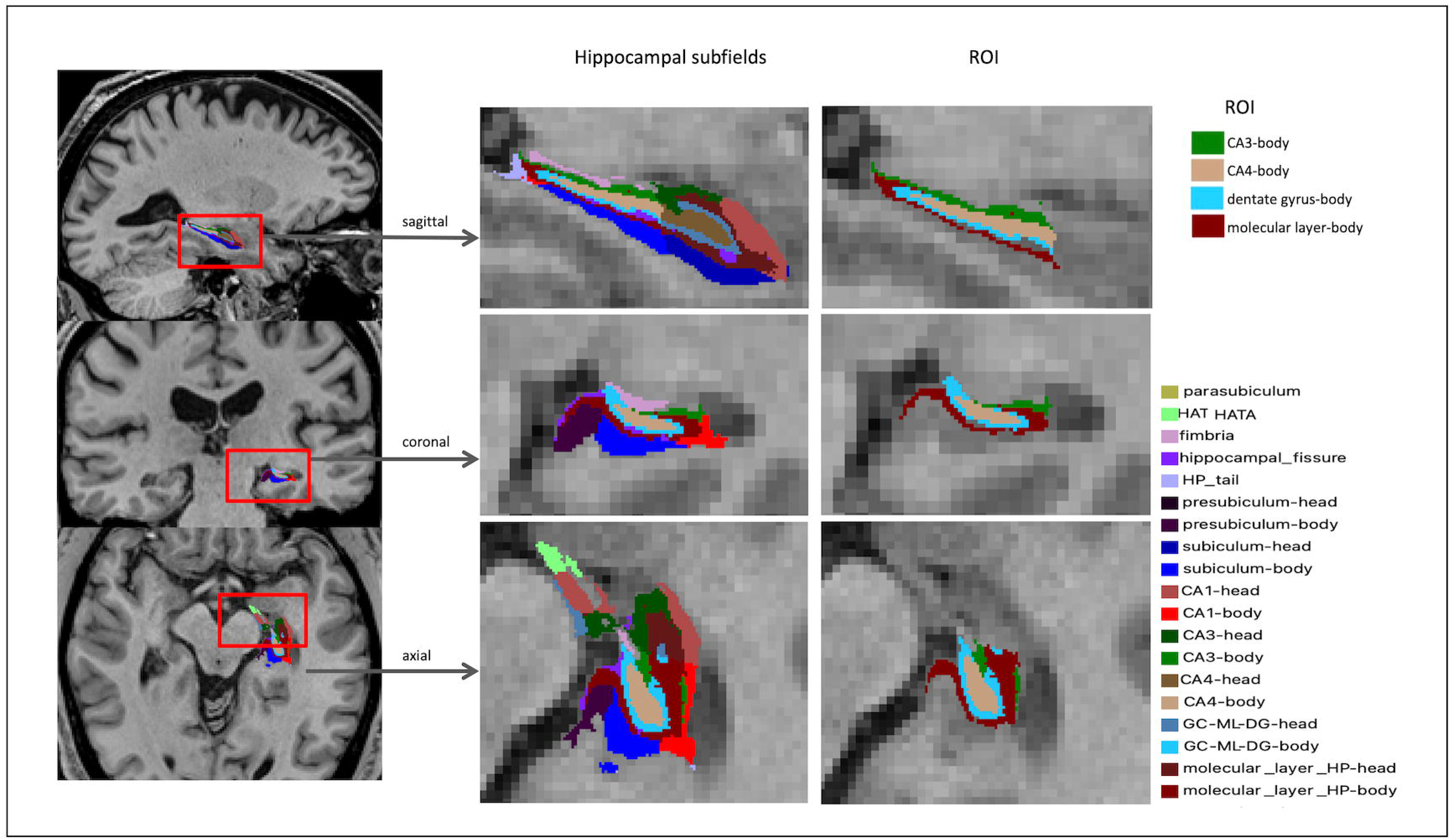
Examples of automatic hippocampal segmentation results by FreeSurfer ver.6 on a T1-weighted MRI in an older adult. Note: HATA = hippocampus-amygdala-transition-area; HP = hippocampus; CA1∼4 = cornu ammonis 1∼4; GC = granule cell layer; ML = molecular layer; DG = dentate gyrus.

Prior to addressing the effect of training on volume in ROI, we first examined whether there was a difference in ROI volume before the intervention between Osc+ and Osc-conditions in each age group. There was no significant difference between conditions in either younger or older adults, *F*(1,94) = 1.30, *p* = .257, *η_p_^2^=* .014, 95% CI [0.000, 0.091] for younger adults and *F*(1,46) = .798, *p* = .376, *η_p_^2^=* .017, 95% CI [0.000, 0.145] for older adults.

To examine our main question of the effect of heart rate oscillation biofeedback training on volume in the LC-innervated-regions hippocampal ROI (i.e., the molecular layer, CA3, CA4 and granule cell layer of dentate gyrus), we performed a three-way mixed ANCOVA (condition x age group x time-point) on ROI volume including condition (Osc+ vs. Osc-) and age group (younger vs. older) as between-subject factors and time-point (pre vs. post) as a within-subject factor while controlling intracranial volume as a covariate (Fig. 3). We found a three-way condition x age group x time-point interaction effect on ROI volume, *F*(1,139) = 6.39, *p* = .013, *η_p_^2^ =* .044, 95% CI [0.019, 0.126]. In younger adults, there was no significant condition x time-point interaction on ROI volume, *F*(1,93) = 0.66, *p* = .419, *η_p_^2^ =* .007, 95% CI [0, 0.075]. In older adults, there was significant condition x time-point interaction on ROI volume, *F*(1,45) = 6.47, *p* = .014, *η_p_^2^=* .126, 95% CI [0.005, 0.307]. Specifically, post-hoc t-tests indicated non-significant opposing trends in the two conditions, with the older Osc+ participants showing increased post-intervention ROI volume compared to pre-intervention ROI volume (*p* = .069) but the older Osc-participants showed decreased post-intervention ROI volume compared to pre-intervention ROI volume (*p* = .079). See Table S4 for mean volumes at pre- and post-intervention time-points for LC-targeted ROI and individual subfields making up the ROI in the left and right hemisphere. To test whether the age-by-condition-by-time-point interaction was modulated by laterality, we performed a four-way mixed ANCOVA (condition x age group x time-point x hemisphere) on ROI volume including condition (Osc+ vs. Osc-) and age group (younger vs. older) as between-subject factors and time-point (pre vs. post) and hemisphere (left vs. right) as a within-subject factor while controlling intracranial volume as a covariate. There was no significant four-way interaction effect, *F*(1,139) = 0.36, *p* = .551, *η_p_^2^ =* .003, 95% CI [0.000, 0.044].

**Fig. 3.**
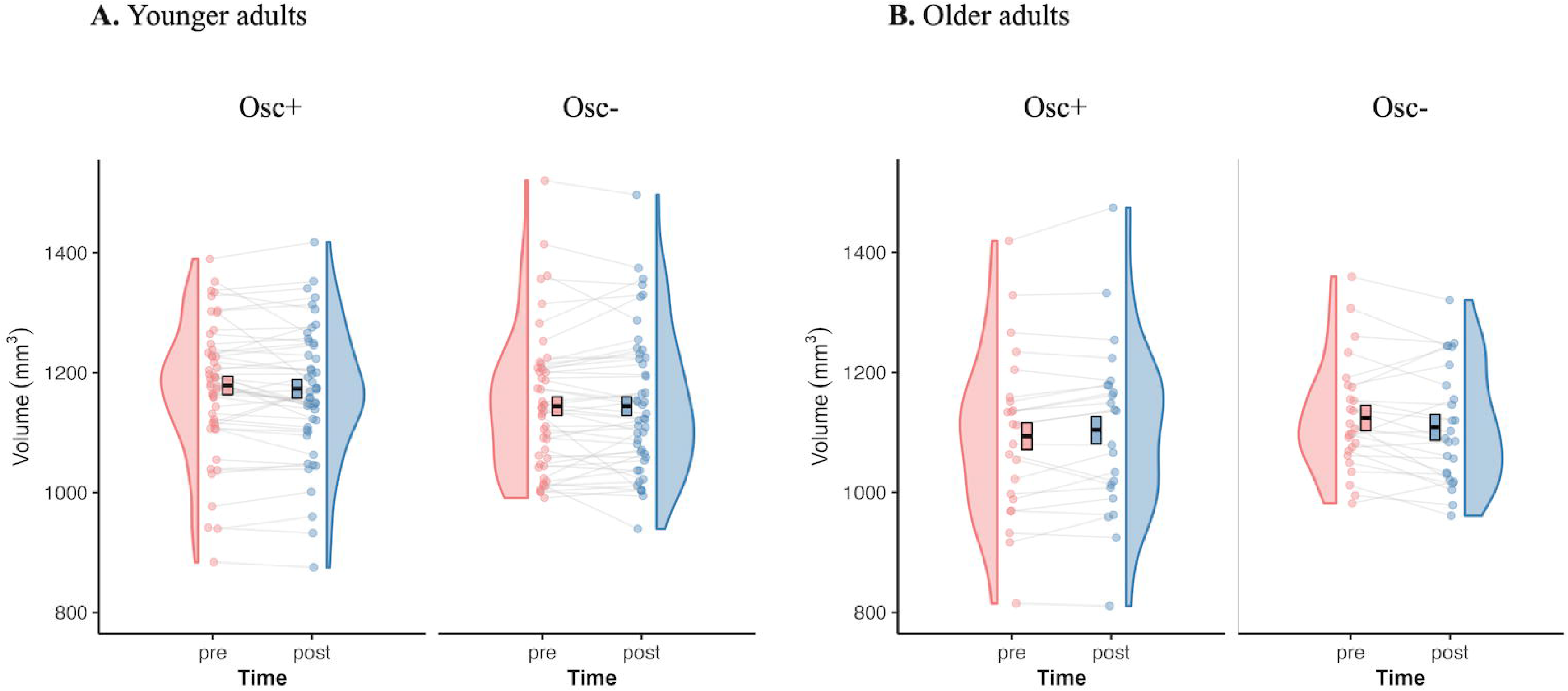
Distribution of hippocampal ROI volume for younger adults (A) and for older adults (B). Mean values were adjusted for intracranial volume as covariate and the error bars reflect the standard error.

We also examined the condition x age group x time-point interaction effect on the whole hippocampus volume controlling intracranial volume and there was no significant interaction, *F*(1,139) = 2.71, *p* = .102, *η_p_^2^* = .019, 95% CI [0, 0.085]. Neither younger nor older adults showed significant differences between pre- and post-intervention scans in volume in the whole hippocampus. To correct the potentially confounding effect across age groups of how many times participants practiced at home, we added the individual total number of training sessions as another covariate in the analysis model. As in our preceding analyses, we found a 3-way condition x age group x time-point interaction effect on ROI volume when controlling for intracranial volume and the number of training sessions, *F*(1,135) = 5.25, *p* = .024, *η_p_^2^* = .037, 95% CI [0, 0.117]. As before, when controlling for ROI volume, in younger adults, there was no significant interaction effect in ROI volume, *F*(1,89) = 0.29, *p* = .594, *η_p_^2^ =* .003, 95% CI [0, 0.063] and in older adults, there was a significant 3-way interaction difference in ROI volume, *F*(1,44) = 7.37, *p* = .009, *η_p_^2^ =* .143, 95% CI [0.009, 0.328]. Additionally we performed correlation analyses to check if the changes in hippocampal ROI volume were associated with the total number of training sessions or total reward. There were no significant correlations between change in hippocampal ROI volume and the total number of training sessions (Osc+: *r* = .145, *p* = .321, Osc-: *r* = .077, *p* = .619 for younger adults and Osc+: *r* = -.275, *p* = .205, Osc-: *r* = .064, *p* = .763 for older adults) and no significant correlations between change in hippocampal volume and the total reward (Osc+: *r* = .004, *p* = .976, Osc-: *r* = -.098, *p* = .514 for younger adults and Osc+: *r* = .379, *p* = .075, Osc-: *r* = -.161, *p* = .441 for older adults). As the older group had a lower proportion of males than the younger group, we checked for effects of sex. Using sex as the 2nd covariate did not change the results as we again found a 3-way condition x age x time-point group interaction effect on ROI volume, *F*(1,138) = 6.38, *p* = .013, *η_p_^2^ =* .044, 95% CI [0.002, 0.127]. Furthermore, when we added sex as another factor neither the interaction effect of sex, condition, age group, and time-point nor the interaction effect of sex, condition, and time-point were significant. The main effects of sex were not significant, either.

About half of the older adults were on cardiac-related medications. Including whether they were or were not on a cardiac-related medication as a factor in the ANCOVA did not lead to a significant main effect of medication group (*p* = .986) nor a condition x time-point x medication group interaction effect (*p* = .958) in the older group.

### 3.3. HRV indices at pre- and post-intervention sessions and during training

We previously reported with this sample that, whereas younger adults showed a significant differential effect of the two interventions on resting log LF-power, older adults did not show any differential effects of the two conditions on any resting HRV metrics (Yoo et al., 2022a). We obtained the same pattern of results with the set of participants included in the current investigation, as well (see Tables 2 and S5). Also, as reported by (Yoo et al., 2022a), during training, there were significant condition differences in total power, log power within the resonance breathing frequency range (within the .063∼.125 Hz range; see Table 2).

**Table 2.**
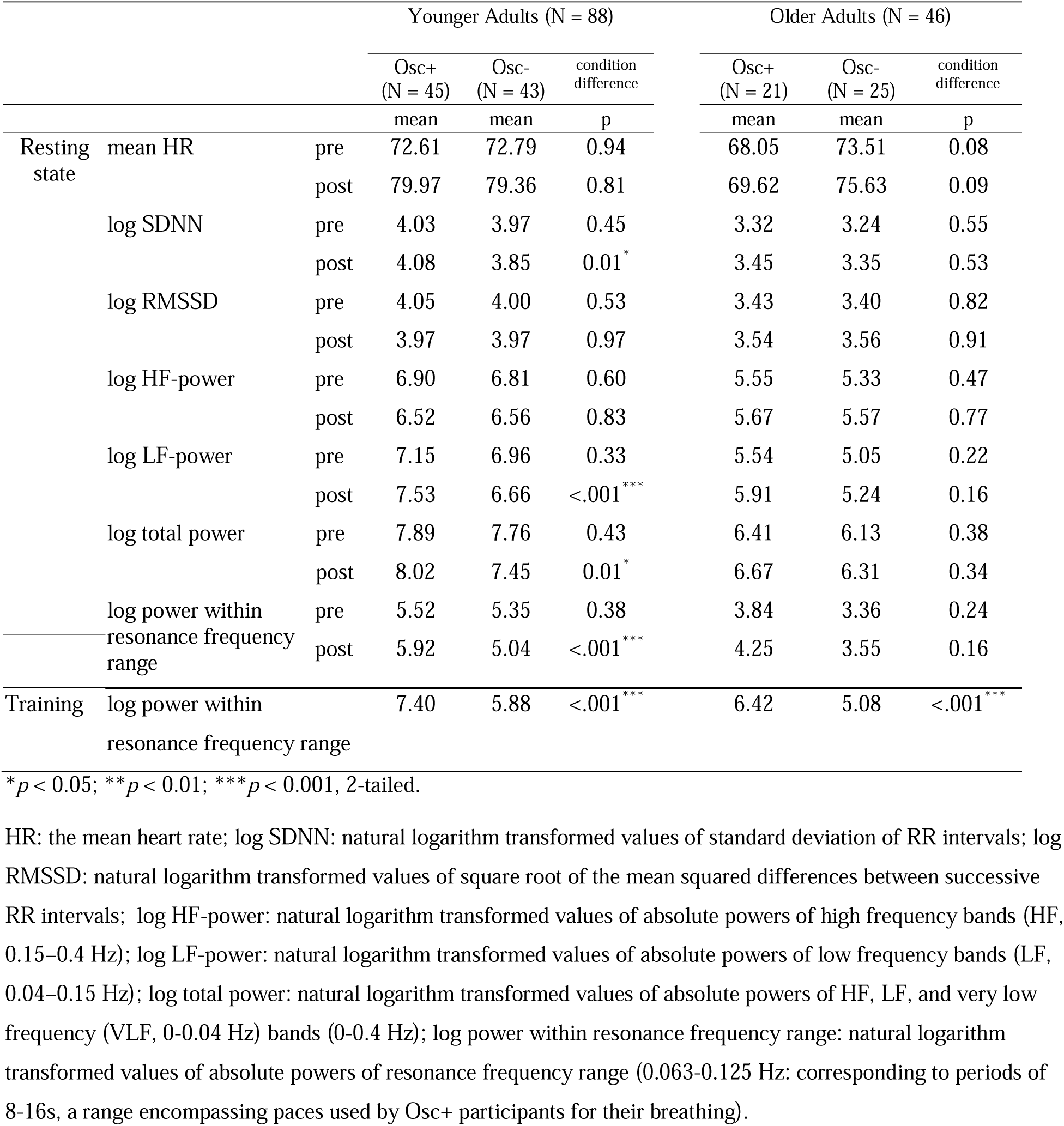
Mean table of HRV indices at pre- and post-intervention sessions and during training (N = 134)

### 3.4. Heart rate oscillatory power and volume changes in the LC-innervated-regions hippocampal ROI

To examine the relationships between heart rate oscillations during practice sessions and hippocampal ROI changes, we first examined the partial correlations between power within the resonance frequency band during training and volume in the LC-innervated-regions hippocampal ROI controlling for pre-intervention ROI volume. We conducted this analysis across conditions because both interventions were framed within the context of meditative states and those Osc-participants who experienced greater relaxation during their home practices may have naturally increased oscillatory power within the frequency range associated with slow breathing. For instance, prior findings (Nesvold et al., 2012) indicate that nondirective meditation increases spectral frequency power within a range overlapping with ours (LF-HRV; 0.04–0.15 Hz). Power within the resonance frequency band during training was positively correlated with ROI volume at the post-intervention scan when controlling for ROI volume at the pre-intervention scan, partial *r* = .428, *p* = .003, 95% CI [.095, .648], but was not significantly correlated for younger adults, partial *r* = -.050, *p* = .648, 95% CI [-.252, .138] (Fig. 4). The correlation *r* value was significantly greater for older adults than for younger adults, *z* = 2.71, *p* = .003. Older adults’ correlation was still significant when controlling for the intervention condition, partial *r* = .307, *p* = .043, 95% CI [-.074, .592], indicating that individual differences in resonance frequency power during training sessions were associated with the hippocampal outcomes even when controlling for the overall condition difference in resonance frequency power. However, when we separated older adults by conditions, neither condition showed significant correlations; *r* = .164, *p* = .490, 95% CI [-.191, 499] for the Osc+ condition and *r* = .385, *p* = .063, 95% CI [-.216, 700] for the Osc-condition., Next, we examined whether power within the resonance frequency band during training mediated the volume change in the hippocampal ROI in older adults. The mediation effect was significant; the path between condition and power within the resonance frequency band during training, a = 1.29, *p* = .0001 and the path between power within the resonance frequency band during training and ROI volume at post, b = 11.24, *p* = .043 (Fig. 5). The results supported a complete mediation model because the direct effect between condition and power within the resonance frequency band during training was not significant, c′ = 10.94, *p* = .385. The results of the mediation analyses suggest that the relationships between condition and volume changes in ROI are fully mediated by power within the resonance frequency band during training.

**Fig 4.**
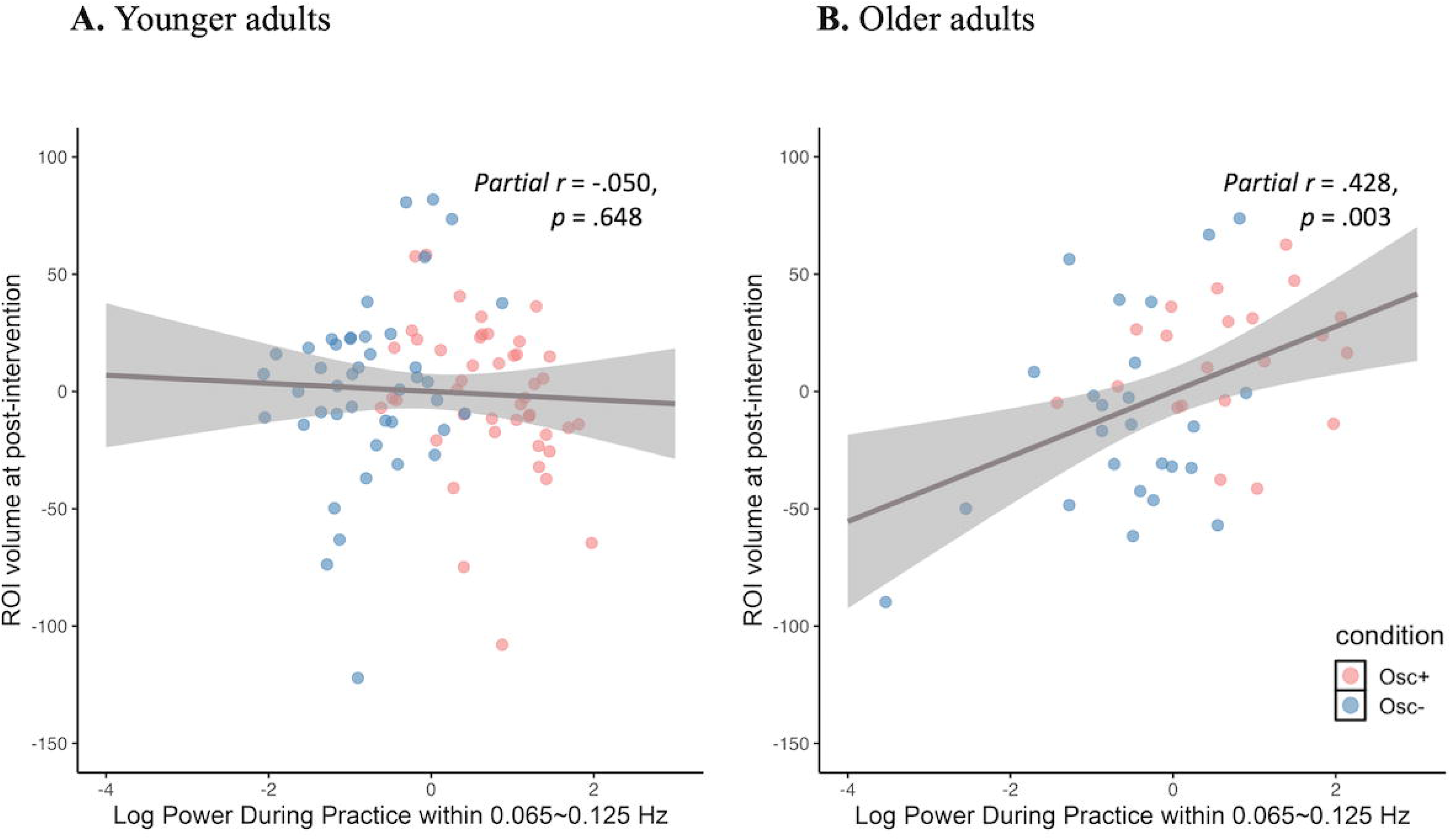
Scatterplot of the partial correlation between heart rate oscillatory activity during training and volume in the LC-innervated-regions hippocampal ROI at post-intervention when controlling for pre-intervention ROI volume. The solid lines represent the fitted regression line, and the shadowed areas represent the 95% confidence interval. Both the X and Y were adjusted for pre-intervention ROI volume in the partial correlation model.

**Fig 5.**
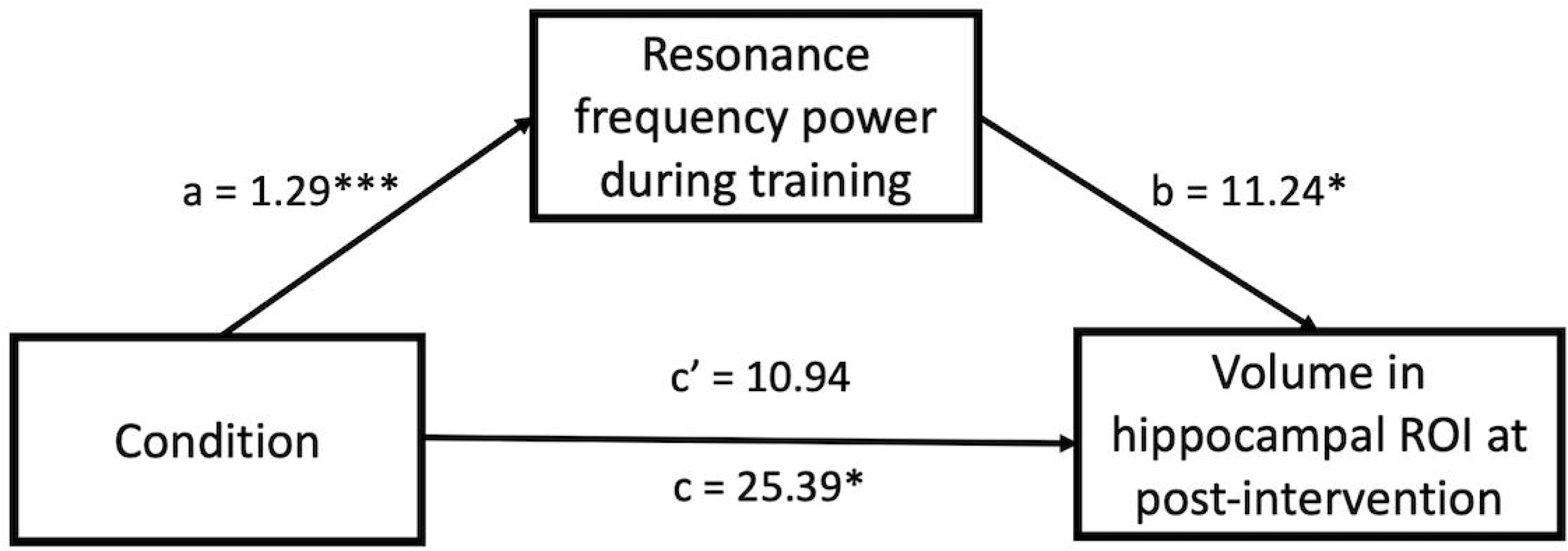
Resonance frequency power mediation model of the relationship between condition and hippocampal ROI volume among older adults. a, b, c, and c’ are expressed as the unstandardized regression coefficients. c represents the total effect and c′ represents the direct effect within the mediation model. \**p* < .05, \*\*\**p* < .001

For comparison, we examined the association between power within the resonance frequency band during training and volume in bilateral whole hippocampus at post-intervention controlling for whole hippocampal volume on the pre-intervention scan. Although associations were in the same direction as for the LC-innervated-regions hippocampal ROI, neither age group showed a significant partial correlation between power within the resonance frequency band during training and volume in bilateral whole hippocampus at post-intervention; in older adults, partial *r* = .183, *p* = .229, 95% CI [-.066, .397], in younger adults, partial *r* = -.105, *p* = .334, 95% CI [-.301, .102]. Given this lack of significant relationship, we did not test a mediation model for the whole hippocampus.

### 3.5. Cerebral blood flow within the LC-innervated-regions hippocampal ROI

Prior to examining CBF within hippocampal ROI, we checked whether there were changes in whole brain gray matter CBF. We applied a three-way ANOVA model (condition x age group x time-point) on whole brain gray matter CBF including condition (Osc+ vs. Osc-) and age group (younger vs. older) as between-subject factors, time-point as within-subject factor. There was no significant 3-way condition x age group x time-point interaction, *F*(1,83) = 0.41, *p* = .525, *η_p_^2^*= .005, 95% CI [0, 0.073]. Next, we examined the same three-way ANOVA (condition x age group x time-point) model on CBF within hippocampal ROI. There was no significant three-way (condition x age group x time-point) interaction, *F*(1,83) = 2.02, *p* = .159, *η_p_^2^* = .024, 95% CI [0, 0.119]. Neither younger or older adults showed significant condition x time-point interactions (younger adults: *p* = .539, older adults: *p* = .128). Thus, it seems unlikely that our findings were due to effects of the intervention on hippocampal CBF.

### 3.6 Exploratory regression analysis to identify predictors

To examine if any variables like demographic characteristics, resting HRV measures, training related measures were related to post-intervention hippocampal ROI volume, we performed exploratory multiple regression analyses for each age group. In younger adults, there were no significant predictors of post-intervention ROI volume except pre-intervention ROI volume. In older adults, power within the resonance frequency band during training was the only significant predictor (*p* = .001) other than pre-intervention ROI volume. Greater power within the resonance frequency band during biofeedback sessions was associated with increases in ROI volume only in older adults. Thus, this exploratory regression result was consistent with the results presented above. The results are summarized in Table S6 in the supplementary information.

## 4. Discussion

In this clinical trial involving random assignment to 5-week interventions involving daily biofeedback to either increase heart rate oscillations while breathing slowly to a pacer (Osc+ condition) or not increase heart rate oscillations while relaxing (Osc-condition), we examined whether the two conditions affected hippocampal volume differently. We found that, in older adults, daily biofeedback sessions modulating heart rate oscillations affected the volume of hippocampal subregions that receive strong noradrenergic input. The Osc+ and Osc-conditions showed significantly different change in hippocampal subregion volume, with the direction of change being increased volume in the Osc+ condition and decreased volume in the Osc-condition. Also, greater average spectral power of heart rate at ∼0.10 Hz (around the breathing frequency used in the Osc+ condition) during biofeedback sessions was associated with increases in volume across these regions in older adults, across conditions. Unlike older adults, younger adults did not show intervention effects on hippocampal volume.

### 4.1. Potential reasons for age differences in the effects of heart rate oscillation interventions

As outlined in the introduction, we hypothesized that the interventions could affect the volume of LC-innervated-regions within the hippocampus via the vagus nerve -> LC -> hippocampus pathway. Considering this pathway, there are at least a couple of reasons that older adults might show greater effects. First, current findings suggest that aging reduces phasic LC activity (Polich and Corey-Bloom, 2005; van Dinteren et al., 2014; Walhovd and Fjell, 2001) while increasing LC tonic activity (Evans et al., 2021). As such, older adults may benefit more than younger adults from an intervention that helps restore phasic input to the LC, or may suffer more from one that suppresses already-weak vagus nerve oscillatory activity. Second, age-related changes in the hippocampal cellular environment (such as changes in neurotrophic factors, mitochondrial activity, synaptic activity, or inflammation (see Kuhn et al., 2018; Mosher and Schaffer, 2018; Smith et al., 2018); may increase the impact of LC noradrenergic input. One age-related change in the hippocampus has particular relevance: animal and human research indicate that hippocampal β-adrenergic receptors increase in number and sensitivity in the prodromal phases of Alzheimer’s disease (Goodman et al., 2021; Kalaria et al., 1989). This could lead older adults to be more sensitive to interventions affecting β-adrenergic activity. Consistent with this possibility, in old mice beta-adrenergic blockers impaired behavior in 3 out of 4 learning and memory tasks, whereas in young mice beta-adrenergic blockers impaired behavior on only one of the tasks (Evans et al., 2021).

### 4.2. Volume changes in this study compared with other interventions and age-related atrophy

A review focusing on structural brain plasticity in adult learning (Lovden et al., 2013) reported that studies using various training protocols like exercise, motor, and cognitive training with acceptable quality had net effects of training on hippocampal volume are in the 2–4% range for 5 days to ∼ 3-month training (Erickson et al., 2011; Lovden et al., 2012; Martensson et al., 2012). If we compute (post-pre)/pre * 100 for our volume values, the Osc+ condition shows a positive 0.96% change. Thus, the increased volume size within the LC-innervated-regions hippocampal ROI in our study is in a similar range compared with the training effects of previous studies.

Hippocampal volume atrophy has not only been found in pathological aging, but also in healthy aging. Also, shrinkage of the hippocampus in healthy adults increased with age (Raz et al., 2005). So we expected, as a default, volume would decrease in the hippocampal ROI and whole hippocampus over time, especially in older adults. The older adults in the Osc-condition showed a 1.35% decrease of volume in the LC-innervated-regions hippocampal ROI, decrements that appear to be greater than the range of normal age-related hippocampal shrinkage (Raz et al., 2005), suggesting that daily practice trying to reduce heart rate oscillatory activity may decrease volume in hippocampal subregions that receive strong noradrenergic signaling.

### 4.3. Potential benefits of phasic vagus nerve activity for the aging hippocampus

Neuronal over-excitation in the hippocampus is a hallmark of early-stage Alzheimer’s disease and aging (Goutagny and Krantic, 2013; Leal and Yassa, 2015). The hippocampus has various properties that make it prone to high excitability that produces seizures (Huberfeld et al., 2015; Mokhothu and Tanaka, 2021). Consistent with the observations of hippocampal hyperexcitation in Alzheimer’s disease, the disease is associated with increased risk of epileptic seizures (Xu et al., 2021). Brief seizures detected using electrocorticogram often lack clinical correlates (Osorio and Frei, 2009). Clinically silent hippocampal seizures may contribute to memory deficits and Alzheimer’s pathological progression (Lam et al., 2017). Thus, hippocampal over-excitation in older adults may accelerate neurodegeneration.

Vagus nerve stimulation is one of the most effective treatments for epilepsy (Ben-Menachem, 2002), raising the possibility that interventions targeting the vagus could benefit older adults starting to experience over-excitation of hippocampal neurons. Via the NTS, vagus nerve activity influences the LC, which in turn modulates activity throughout much of the brain, including the hippocampus. In order for vagus nerve stimulation to be effective for epilepsy treatment, the locus coeruleus needs to be intact and functioning (Fornai et al., 2011; Krahl et al., 1998). Furthermore, studies that have demonstrated benefits of vagus nerve stimulation on hippocampal plasticity and/or memory performance have employed bursts of vagus nerve stimulation rather than constant stimulation (Biggio et al., 2009; Clark et al., 1998; Sanders et al., 2019). This type of vagus nerve stimulation increases phasic LC activity (Hulsey et al., 2017). Further evidence that intermittent vagus nerve stimulation increases phasic LC activity comes from a study examining P3 event-related potentials in response to oddball stimuli in epileptic patients (De Taeye et al., 2014). Epileptic patients who had experienced more than a 50% reduction in seizures due to vagus nerve stimulation showed greater P3 responses (an indicator of phasic LC activity; Vazey et al., 2018) shortly after vagus nerve stimulation than after sham stimulation. Slow paced breathing provides a naturally oscillating set of input signals to the LC via the vagus nerve, which may alternate between inhibiting LC activity (thereby reducing stress-like patterns of high tonic LC activity) and exciting LC activity (promoting phasic bursts of activity). Direct stimulation of the LC in a rat model of prodromal Alzheimer’s disease has demonstrated that phasic LC activity benefits memory compared with tonic activity (Omoluabi et al., 2021). Thus, slow-paced breathing may benefit the aging hippocampus by promoting phasic LC activity. Conversely, attempting to keep one’s heart rate steady and reduce oscillatory activity may promote tonic LC activity that is detrimental to the aging hippocampus.

### 4.4. Potential implications for memory and cognition

Many studies have indicated that hippocampal volume can predict performance on a variety of cognitive tasks in healthy controls (Convit et al., 2003; Hackert et al., 2002; Rosen et al., 2003; Van Petten, 2004), although correlations are not always seen (e.g., MacLullich et al., 2002; Maguire et al., 2000; Marquis et al., 2002). In addition, hippocampal volume has been suggested as a predictive measure of deterioration of memory function in a variety of neurodegenerative diseases including Alzheimer disease (AD) (den Heijer et al., 2006; Dubois et al., 2014) and Parkinson’s disease (Brück et al., 2004). Future studies of biofeedback training to modulate heart rate oscillations should examine the relationship between hippocampal volume change, and change in performance on cognitive tasks in healthy older adults, mild cognitive impairment (MCI), and AD groups.

### 4.5. Relationships between meditation and hippocampal volume in older adults may relate to heart rate oscillatory activity during these practices

Previous studies have shown that older adults who practice Tai Chi or meditate regularly tend to have larger hippocampal volume than those who do not meditate (Fox et al., 2014; Holzel et al., 2008; Liu et al., 2019; Luders et al., 2013; Luders et al., 2009; Yue et al., 2020). Tai Chi practitioners over age 60 show greater hippocampal grey matter density than do brisk walkers over age 60 (Yue et al., 2020), suggesting the Tai Chi effects go beyond the aerobic benefits of the activity. Increases in heart rate oscillations occur in meditators practicing Qigong, Kundalini yoga, or Zazen (Lehrer et al., 1999; Peng et al., 2004; Peng et al., 1999) or in people reciting either the rosary or a typical yoga mantra (Bernardi et al., 2001). Tai Chi also increases abdominal breathing and heart rate oscillatory activity (Wei et al., 2016). During these meditative practices, breathing slows down and drives large heart rate oscillations at the breathing frequency (Bernardi et al., 2001; Lehrer et al., 1999; Peng et al., 2004). Thus, one possibility is that meditative practices that increase heart rate oscillations may help older adults avoid hippocampal atrophy.

### 4.6. Limitations and alternative mechanisms

A limitation of these analyses was that we were limited to the available 1□×□1□×□1□mm^3^ resolution T1-weighted MRI images. Unlike high resolution T2-weighted images, 1□×□1□×□1□mm^3^ T1-weighted images do not allow clear visualization of the stratum radiatum lacunosum moleculare (SRLM). Due to this limitation, we avoided analyzing subfield data from the hippocampal head where the SRLM is needed to visualize the hippocampal digitations (Wisse et al., 2021). We focused instead on the hippocampal body, which has a less complex structure for segmentation. In addition, to reduce the impact of idiosyncratic errors in any one subfield, we combined four adjacent LC-targeted subregions from the hippocampal body into one larger ROI. Reliability was high across hippocampal subregions across the two scans despite the intervening intervention (Table S3) and each of the separate subfields comprising the LC-targeted ROI showed a consistent pattern in which volume shows relatively more positive change in the Osc+ than in the Osc-condition for older adults (Table S4).

It is also important to note that there are alternative pathways other than via LC modulation through which the daily practice of heart rate oscillation biofeedback could influence hippocampal volume. For instance, the hippocampus is especially susceptible to impaired cerebrovascular dynamics (Kril et al., 2002; Raz et al., 2007), which may have been affected by the intervention. For instance, if the intervention increased blood flow to the hippocampus, this could potentially influence hippocampal volume (e.g., Fierstra et al., 2010). For this reason, we checked whether there were changes in blood flow and whether they could account for our volumetric findings. Average cerebral blood flow across the gray matter regions in the brain did not show any significant intervention effects. There was no significant three-way (condition x age group x time-point) interaction on CBF within hippocampal ROI.

### 4.7. Conclusions

In conclusion, our results showed that daily practice of heart rate oscillation biofeedback can affect the volume in bilateral hippocampal subregions innervated by LC (CA3 body, CA4 body, dentate gyrus, molecular layer) in older adults. Among older adults, there were significant differences across conditions in hippocampal ROI, with the Osc+ condition showing relatively increased volume and Osc-showing relatively decreased volume. Furthermore, only in older adults, spectral power of heart rate within the resonance frequency band during training was positively correlated with volume change in hippocampal subregions innervated by LC. These findings provide novel evidence for causal pathways between heart rate oscillatory activity and the aging hippocampus.

## Data Availability

All data produced are available online at OpenNeuro (https://openneuro.org/datasets/ds003823).

## Data/code availability

The empirical data used for this paper are available in the public repository, OpenNeuro, “HRV-ER” (https://openneuro.org/datasets/ds003823). The scripts used for hippocampal volume analysis using Freesurfer 6.0 and segmentation tool of hippocampal subfields and nuclei of the amygdala are freely available online (https://github.com/EmotionCognitionLab/HRV-ER_hippocampus) and codes for other statistical analyses are available upon request from the corresponding author.

## Acknowledgments

This study was supported by NIH R01AG057184 (PI Mather). We thank study participants for their contribution.We thank our research assistants for their help with data collection: Paul Choi, Heekyung Rachael Kim, Seungyeon Lee, Alexandra Haydinger, Lauren Thompson, Gabriel Shih, Divya Suri, Sophia Ling, Akanksha Jain, Linette Bagtas, Michelle Wong, Kathryn Cassutt, Collin Amano, Yong Zhang.

## CRediT authorship contribution statement

The authors made the following contributions. HY: Conceptualization, Data curation, Formal analysis, Investigation, Methodology, Software, Validation, Visualization, Writing - original draft; KN: Conceptualization, Data curation, Investigation, Writing - review & editing, Project administration; SD: Formal analysis, Investigation, Methodology, Writing; JM: Conceptualization, Data curation, Investigation; CCho: Conceptualization, Data curation, Investigation, Resources, Project administration; JFT, PL, & CChang: Conceptualization, Writing - review & editing; MM: Conceptualization, Funding acquisition, Resources, Project administration, Supervision, Writing - review & editing.

