## Supplementary Materials for "Daily biofeedback to modulate heart rate oscillations affects structural volume in hippocampal subregions targeted by the locus coeruleus in older adults but not younger adults"

### S1. Supplementary Methods

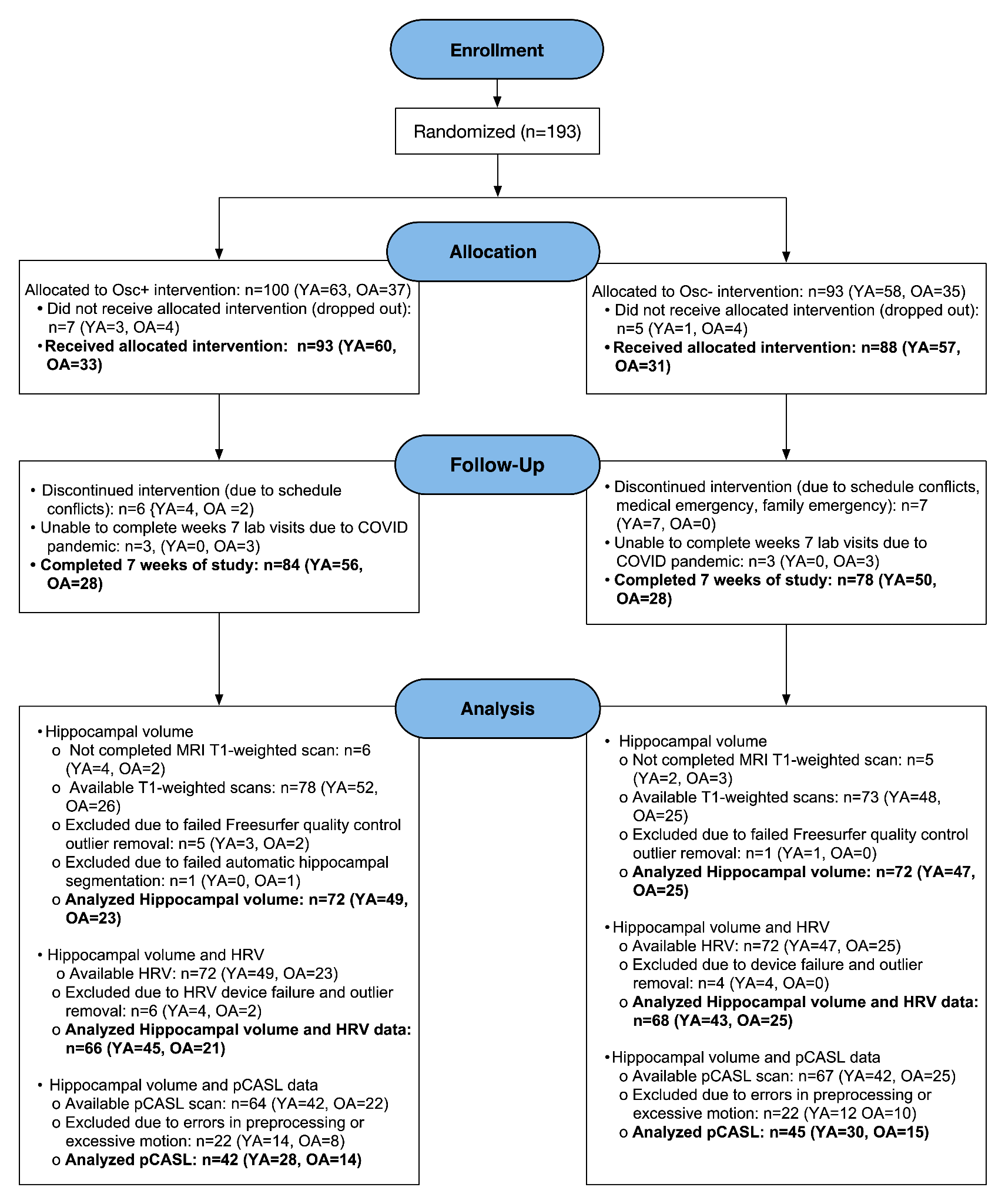

**Figure S1.** Numbers of participants in each intervention condition, how many participants completed each measure, and how many were included vs. excluded in each analysis.

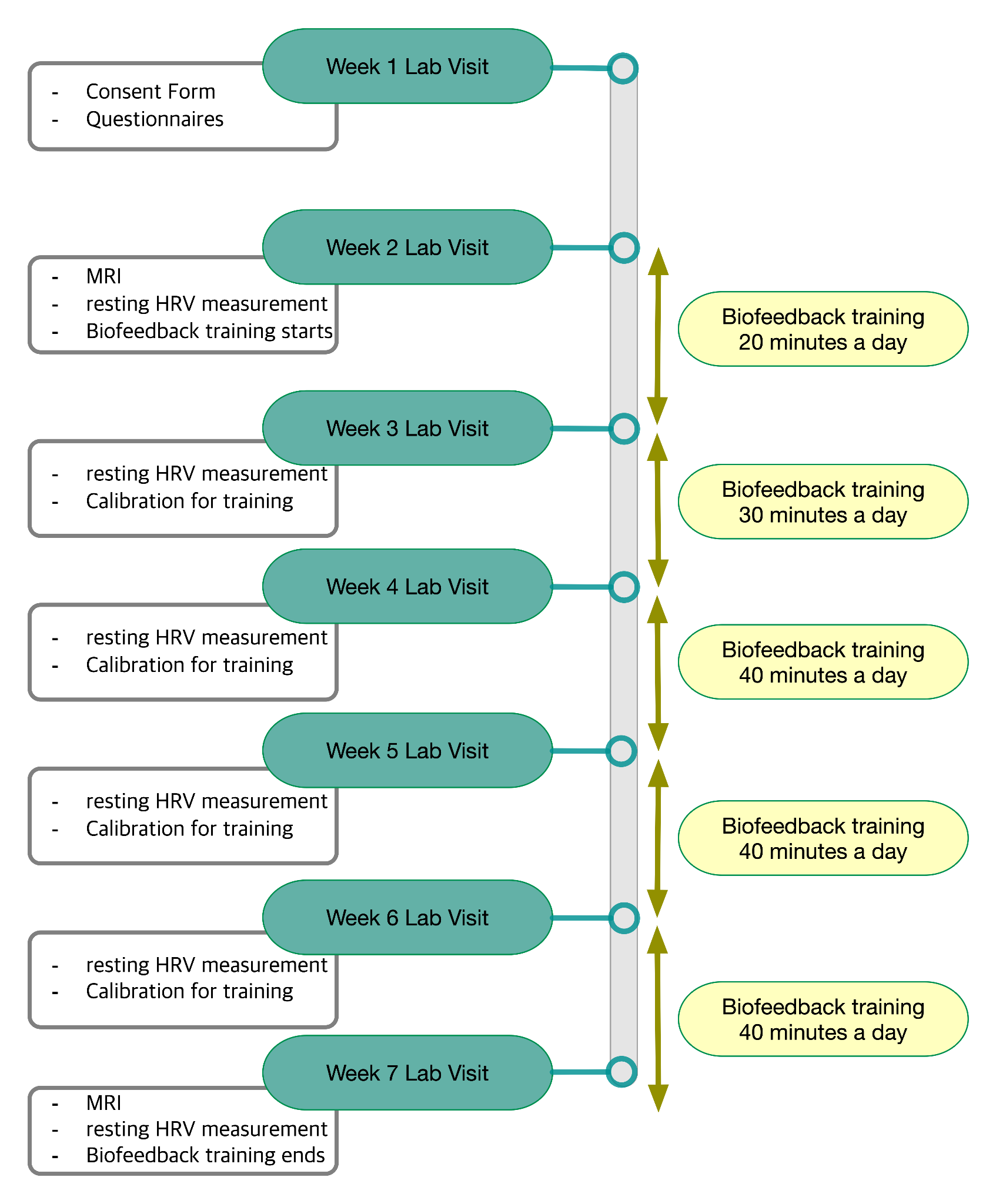

**Figure S2.** Study 7-week schedule.

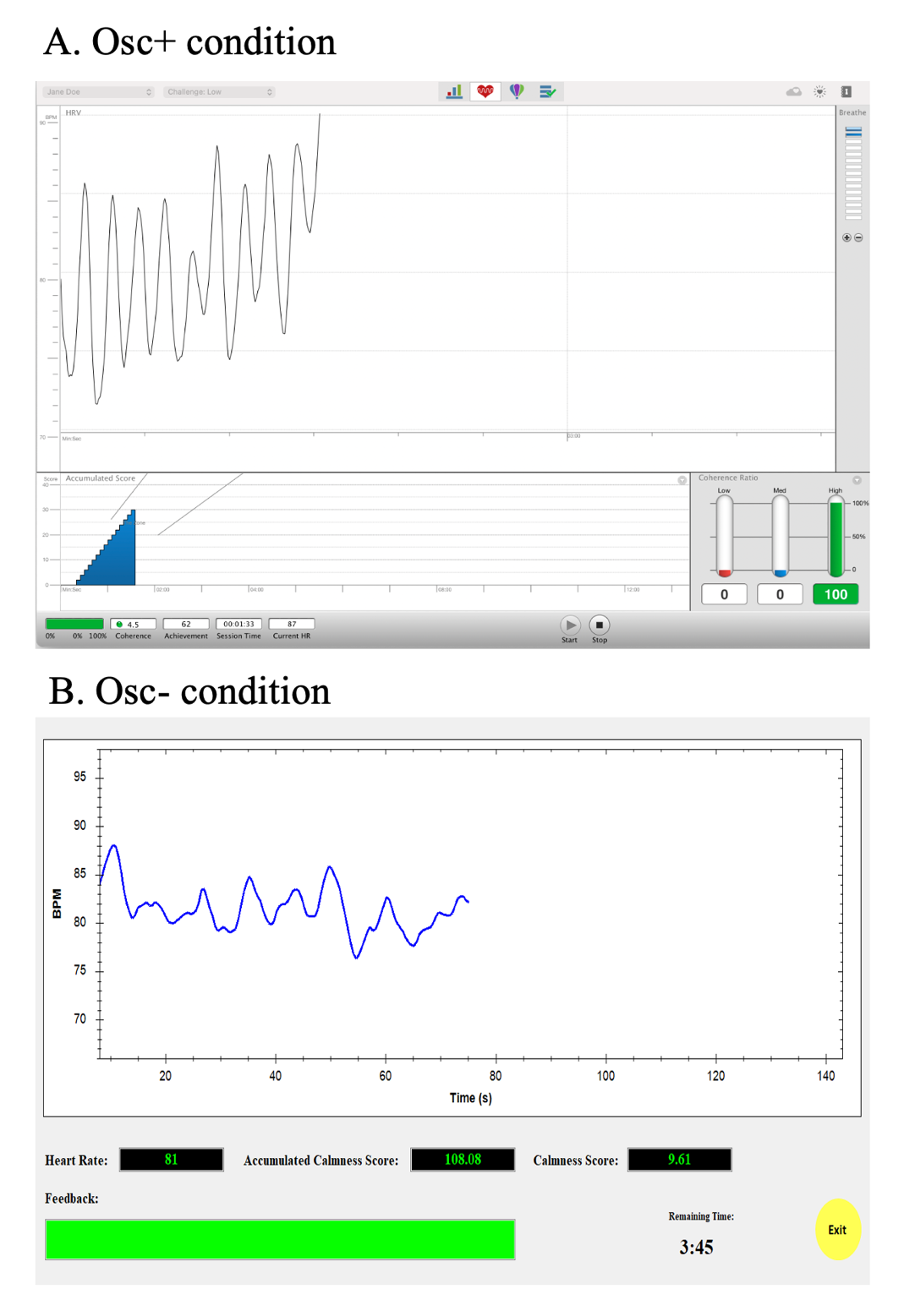

**Figure S3.** Training display for Osc+ (A) and Osc- (B) condition. The Osc+ condition used emWave software (HeartMath®Institute, 2020) to give positive feedback for heart rate oscillations corresponding with slow breathing frequencies whereas the Osc- condition used custom software to show positive feedback when heart rate oscillatory activity was low.

S1.1. Procedure

##### S1.1.1. Overview of 7-week Protocol Schedule

The study protocol involved seven weekly lab visits and five weeks of home biofeedback training. The first lab visit involved the non-MRI baseline measurements, including a number of questionnaires. The second lab visit involved the baseline MRI session, then followed by the first biofeedback calibration and training session. Each lab calibration session started with a 5-min baseline rest period followed by several 5-min intervals practicing different strategies to find the best condition. After calibration sessions were completed, participants were notified which strategy was the best and were requested to practice the best condition at home for 10 min twice a day for the 1st training week (between the 1st week visit and the 2nd week visit), 15 min twice a day for the 2nd training week (between the 2nd week visit and the 3rd week visit), and 20 min twice a day for the last weeks (between the 3rd week visit and the 7th week visit).

The weekly lab visits (except for weeks with MRI sessions) were run in small groups of participants from the same condition in which participants shared their experiences and tips about biofeedback training with other participants, while 1-2 researchers facilitated the discussion. Outside the lab, participants used a customized social app to communicate with other members of their group and researchers about their progress on daily biofeedback training. For instance, participants gave each other 'thumbs-up' or smiling face emojis when they completed training for the day, and researchers sent participants a friendly reminder to complete home training when they were falling behind. The week-6 lab visit repeated the assessments from the first lab visit. The final (7th) lab visit repeated the baseline MRI session scans in the same order, followed by a few additional scans not reported here (see Yoo et al., 2022 for details ).

##### S1.1.2. Biofeedback Training for the Osc+ condition

During all practice sessions, participants wore an ear sensor to measure their pulse. They viewed real-time heart rate biofeedback while breathing in through the nose and out through the mouth in synchrony with the emWave pacer. The emWave software (HeartMath®Institute, 2020) provided a summary ‘coherence’ score for participants that was calculated as peak power/(total power - peak power), with peak power determined by finding the highest peak within the range of .04 - .26 Hz and calculating the integral of the window .015 Hz above and below this highest peak, and total power computed for the .0033 - .4 Hz range.

During the second lab visit, we introduced participants to the device and identified the resonance frequency for each participant during five minutes of paced breathing at 6, 6.5, 5.5, 5 and finally 4.5 breaths/min (Lehrer et al., 2013). After all 5-min breathing segments were complete, we computed various aspects of the oscillatory dynamics for each breathing pace using Kubios HRV Premium 3.1 software (Tarvainen et al., 2014) and estimated which breathing pace best approximated the resonance frequency by assessing which one had the most of the following characteristics: highest low frequency (LF) power, the highest maximum LF amplitude peak on the spectral graph, highest peak-to-trough amplitude, cleanest and highest-amplitude LF peak, highest coherence score and highest the root mean squared successive differences (RMSSD). Participants were then instructed to train at home with the pacer set to their identified resonance frequency and to try to maximize their coherence scores.

During the third visit, they were asked to complete three 5-min paced breathing segments: the best condition from the last week’s visit, half breath per minute faster and half breath slower than the best condition. They were then instructed to train the following week at the pace that best approximated the resonance frequency based on the characteristics listed above. In subsequent weekly visits, during 5-min training segments, they were asked to try out abdominal breathing and inhaling through nose/exhaling through pursed lips as well as other strategies of their choice.

##### S1.1.3. Biofeedback Training for the Osc- condition

The same biofeedback ear sensor was used in this condition as in the Osc+ condition. However, we created custom software to display a different set of feedback to the Osc- participants. During each Osc- training session, a ‘calmness’ score was provided as feedback to the participants instead of the coherence score. The calmness score was calculated by multiplying the coherence score that would have been displayed in the Osc+ condition by -1 adding 10 (an ‘anti-coherence’ score). Thus, participants got more positive feedback (higher calmness scores) when their heart rate oscillatory activity in the 0.04 – 0.26 Hz range was low.

Participants also received a point adjustment that gave a penalty when heart rate was the lowest it had been in the past 15 s. Specifically, every 5 s, a local maximum IBI was set based on the maximum IBI from the last 15 s. If, at that point, the participant’s current IBI was longer than this local maximum, the calmness score displayed for the next 5 s was the anti-coherence score - 2. Naturally, most of the time, current IBI was lower than the local maximum, and in those cases, the calmness score was the anti-coherence score +1. Thus, there was a penalty in their calmness score for moments when their heart rate was slower than it had been in any of the past 15 s. However, average heart rate during biofeedback sessions did not differ significantly across conditions. Thus, this additional feedback appeared to have had little impact on heart rate.

During the initial calibration session at the end of the second lab visit, each participant was introduced to the device and feedback and was asked to come up with five strategies to lower heart rate and heart rate oscillations. The participant was instructed to wear the ear sensor and view real-time heart rate biofeedback while they tried each strategy for five minutes. We analyzed the data in Kubios and identified the best strategy as the one that had the most of the following characteristics: lowest LF power, the minimum LF amplitude peak on the spectral graph, lowest peak trough amplitude, multiple and lowest-amplitude LF peak, highest calmness score and lowest RMSSD. Participants were then instructed to use this strategy to try to maximize their calmness scores in their home training sessions.

On the third visit, they were asked to select three strategies and try each out in a 5-min session. The strategy identified as best (based on the same characteristics used in the initial calibration session) was selected as the one to focus on during home training. In subsequent weekly visits, during 5-min training segments, they were asked to try out strategies of their choice.

##### S1.1.4. Rewards for Performance

In addition to receiving compensation of $15 per hour for each lab visit, participants were eligible to receive rewards based on individual and group performance. For individual performance rewards, each week participants had the opportunity to earn $2 for each instance (up to a maximum of 10) they exceeded their assigned target score (target scores were assigned each week and were the average of the top 10 scores earned from the previous week’s training sessions plus 0.3). Group performance rewards were earned when members of a participant's group completed a minimum of 80% of their assigned biofeedback training minutes. For example, if a participant completed 100% of their training, they received an additional $3 for each group member who also completed 100% of their training. If a participant completed 80% of their training, they received an additional $2 for each group member who also completed at least 80% of their training. Rewards were calculated weekly, and participants received weekly updates on their earnings at their lab visits. There was no significant condition difference or significant condition x age group interaction in total rewards.

**S2. Supplementary Results**

**Table S1.** Categories of medication reported across age groups and conditions

|  | Younger adults | | | |  | Older adults | | | |
| --- | --- | --- | --- | --- | --- | --- | --- | --- | --- |
|  | N |  | Osc+ | Osc- |  | N |  | Osc+ | Osc- |
| No medication | 77 |  | 36 | 34 |  | 13 |  | 6 | 7 |
| Medication (≧1 type) | 19 |  | 9 | 9 |  | 35 |  | 15 | 18 |
| Subcategory |  |  |  |  |  |  |  |  |  |
| Antihistamine | 1 |  | 1 |  |  | 2 |  | 1 | 1 |
| Cardiac medication |  |  |  |  |  | 24 |  | 13 | 11 |
| Vitamin/herbal supplement | 1 |  |  | 1 |  | 10 |  | 5 | 5 |
| Pain medication | 3 |  | 2 | 1 |  | 10 |  | 3 | 7 |
| Psychiatric medication | 3 |  |  | 3 |  | 11 |  | 6 | 5 |
| Sedative/hypnotic | 1 |  | 1 |  |  | 3 |  | 1 | 2 |
| Hormones | 12 |  | 6 | 6 |  | 7 |  | 2 | 5 |
| Other | 1 |  | 1 |  |  | 10 |  | 5 | 5 |

**Table S2.** Mean and standard deviation of total rewards. Unit = US dollar

|  | Younger Adults | Older Adults |
| --- | --- | --- |
| Osc+ | 298.09 (32.54) | 447.85 (58.57) |
| Osc- | 293.82 (40.62) | 425.16 (55.52) |
| Condition main effect p-value | *p* =.570 | *p* = .175 |

**Table S3.** Test-retest reliability (*r*) for hippocampal subregions

|  | region | Left | Right |
| --- | --- | --- | --- |
| Subregion | Hippocampal tail | .964** | .961** |
|  | subiculum-body | .960** | .965** |
|  | CA1-body | .961** | .968** |
|  | subiculum-head | .963** | .971** |
|  | hippocampal-fissure | .857** | .866** |
|  | presubiculum-head | .934** | .957** |
|  | CA1-head | .982** | .980** |
|  | presubiculum-body | .966** | .967** |
|  | parasubiculum | .923** | .945** |
|  | molecular layer-head | .979** | .975** |
|  | molecular layer-body | .948** | .949** |
|  | dentate gyrus-head | .967** | .968** |
|  | CA3-body | .963** | .957** |
|  | dentate gyrus-body | .927** | .921** |
|  | CA4-head | .954** | .958** |
|  | CA4-body | .923** | .925** |
|  | fimbria | .919** | .919** |
|  | CA3-head | .977** | .972** |
|  | HATA | .923** | .933** |
| ROI | 4 subregions innervated by LC (CA3 body, CA4 body, dentate gyrus, molecular layer) | .942** | .940** |
| Whole | Whole hippocampal-body | .957** | .963** |
|  | Whole hippocampal-head | .983** | .977** |
|  | Whole hippocampus | .982** | .979** |

**p* < .05, ***p* < .01

**Table S4.** Mean pre- and post-intervention volumes for individual subfields making up the LC-targeted ROI

|  | Region |  | Younger Adults | | |  | Older Adults | | |  |  |  |
| --- | --- | --- | --- | --- | --- | --- | --- | --- | --- | --- | --- | --- |
|  |  |  | Osc+ | Osc- | Condition x time-point interaction |  | Osc+ | Osc- | Condition x time-point interaction |  | Condition x age group x time-point interaction | |
|  |  |  | mean | mean | *p* |  | mean | mean | *p* |  | *p* | *η_p_^2^* |
| Left | ROI total | pre | 570.22 | 548.69 | .42 |  | 522.11 | 527.58 | .19 |  | .098 | .020 |
|  |  | post | 567.68 | 548.93 |  |  | 523.95 | 519.41 |  |  |  |  |
|  | CA3 body | pre | 95.33 | 88.77 | .47 |  | 86.13 | 90.11 | .27 |  | .170 | .014 |
|  |  | post | 94.15 | 88.19 |  |  | 86.73 | 88.74 |  |  |  |  |
|  | CA4 body | pre | 120.22 | 115.52 | .22 |  | 109.70 | 111.92 | .10 |  | .025* | .036 |
|  |  | post | 119.92 | 116.31 |  |  | 110.51 | 109.65 |  |  |  |  |
|  | dentate gyrus-body | pre | 130.81 | 126.60 | .23 |  | 122.19 | 121.77 | .14 |  | .042* | .029 |
|  |  | post | 130.40 | 127.24 |  |  | 122.93 | 119.80 |  |  |  |  |
|  | molecular layer-body | pre | 223.86 | 217.81 | .90 |  | 204.10 | 203.77 | .37 |  | .422 | .005 |
|  |  | post | 223.21 | 217.19 |  |  | 203.78 | 201.22 |  |  |  |  |
| Right | ROI total | pre | 608.45 | 595.13 | .56 |  | 571.35 | 596.40 | .03* |  | .025* | .036 |
|  |  | post | 605.49 | 594.99 |  |  | 579.90 | 589.06 |  |  |  |  |
|  | CA3 body | pre | 110.77 | 106.10 | .29 |  | 105.44 | 115.61 | .01* |  | .009* | .048 |
|  |  | post | 109.53 | 106.36 |  |  | 107.72 | 113.49 |  |  |  |  |
|  | CA4 body | pre | 125.68 | 123.49 | .48 |  | 118.94 | 121.05 | .07 |  | .039* | .030 |
|  |  | post | 124.89 | 123.47 |  |  | 120.63 | 119.36 |  |  |  |  |
|  | dentate gyrus-body | pre | 134.65 | 133.16 | .37 |  | 131.30 | 131.54 | .11 |  | .052 | .027 |
|  |  | post | 133.87 | 133.36 |  |  | 133.11 | 130.44 |  |  |  |  |
|  | molecular layer-body | pre | 237.35 | 232.39 | .78 |  | 215.66 | 228.22 | .04* |  | .099 | .019 |
|  |  | post | 237.21 | 231.81 |  |  | 218.43 | 225.77 |  |  |  |  |
| Bi-  lateral | ROI total | pre | 1178.67 | 1143.82 | .42 |  | 1093.46 | 1123.98 | .01* |  | .013* | .044 |
|  |  | post | 1173.17 | 1143.92 |  |  | 1103.84 | 1108.47 |  |  |  |  |

**p* < 0.05; ***p* < 0.01; ****p* < 0.001, 2-tailed. Condition x time-point interaction effects were statistically tested using two-way mixed ANCOVA on volume including condition (OSC+ vs. OSC-) as between-subject factor, time-point( pre vs. post) as within-subject factor, and intracranial volume as a covariate. Condition x age group x time-point interaction effects were statistically tested using a three-way mixed ANCOVA on volume including intracranial volume as a covariate.

**Table S5.** P values and partial eta-squared values from the three-way ANOVA and two-way ANOVAs with each resting state HRV measure as dependent variable.

|  |  |  |  |  | Younger Adults (N = 88) | |  | Older Adults (N = 46) | |
| --- | --- | --- | --- | --- | --- | --- | --- | --- | --- |
|  |  | Condition X age group X time-point | |  | Condition X time-point | |  | Condition X time-point | |
|  |  | *p* | *η_p_^2^* |  | *p* | *η_p_^2^* |  | *p* | *η_p_^2^* |
| Resting state | mean HR | .707 | .001 |  | .743 | .001 |  | .755 | .002 |
|  | log SDNN | .297 | .008 |  | .052 | .043 |  | .900 | .000 |
|  | log RMSSD | .968 | .000 |  | .580 | .004 |  | .743 | .002 |
|  | log HF-power | .962 | .000 |  | .492 | .006 |  | .677 | .004 |
|  | log LF-power | .247 | .010 |  | **.007**** | .081 |  | .633 | .005 |
|  | log total power | .304 | .008 |  | **.031*** | .053 |  | .783 | .002 |

**p* < 0.05; ***p* < 0.01; ****p* < 0.001, 2-tailed.

**Table S6.** Multiple linear regression analysis using hippocampal ROI volume at post-intervention as dependent variable

| Group | Variables | Unstandardized  Coefficients | | Standardized Coefficients | t | Sig. | 95.0% Confidence Interval for B | |
| --- | --- | --- | --- | --- | --- | --- | --- | --- |
|  |  | B | Std. Error | Beta |  |  | Lower Bound | Upper Bound |
| Younger adults | (Constant) | 172.947 | 90.841 |  | 1.904 | .060 | -7.731 | 353.625 |
|  | Age | .705 | 2.456 | .017 | .287 | .775 | -4.180 | 5.590 |
|  | Gender | -2.609 | 8.039 | -.011 | -.325 | .746 | -18.598 | 13.381 |
|  | Education | -2.491 | 3.175 | -.047 | -.785 | .435 | -8.805 | 3.824 |
|  | Total number of training sessions | .126 | .185 | .023 | .682 | .497 | -.243 | .495 |
|  | mean HR | -.622 | .443 | -.053 | -1.405 | .164 | -1.503 | .259 |
|  | Log SDNN | -11.513 | 21.931 | -.041 | -.525 | .601 | -55.133 | 32.108 |
|  | Log RMSSD | -6.637 | 21.296 | -.025 | -.312 | .756 | -48.995 | 35.720 |
|  | Log power within  resonance frequency range | -.175 | 3.904 | -.002 | -.045 | .964 | -7.940 | 7.589 |
|  | Hippocampal ROI volume at pre | .966 | .034 | .963 | 28.380 | <.001*** | .898 | 1.034 |
| Older adults | (Constant) | 69.298 | 134.333 |  | .516 | .609 | -202.646 | 341.242 |
|  | Age | -.386 | .950 | -.022 | -.406 | .687 | -2.309 | 1.537 |
|  | Gender | 7.848 | 12.087 | .030 | .649 | .520 | -16.620 | 32.317 |
|  | Education | .308 | 2.644 | .006 | .116 | .908 | -5.045 | 5.661 |
|  | Total number of training sessions | -.473 | .356 | -.068 | -1.329 | .192 | -1.194 | .248 |
|  | mean HR | -.753 | .633 | -.063 | -1.190 | .242 | -2.035 | .529 |
|  | Log SDNN | -17.303 | 30.508 | -.079 | -.567 | .574 | -79.064 | 44.457 |
|  | Log RMSSD | -2.686 | 29.623 | -.012 | -.091 | .928 | -62.654 | 57.283 |
|  | log power within  resonance frequency range | 15.698 | 4.841 | .161 | 3.243 | .002** | 5.897 | 25.498 |
|  | Hippocampal ROI volume at pre | 1.011 | .046 | .981 | 21.834 | <.001*** | .917 | 1.105 |

**p* < 0.05; ***p* < 0.01; ****p* < 0.001. Younger adults: *R* = .957, *R^2^* = .915; Older adults: *R* = .965, *R^2^* = .931
